# The Neurocognome in Childhood: Relevance of Neurocognitive Network Organization

**DOI:** 10.1101/2023.05.30.23290689

**Authors:** M. Königs, C.C. Kooper, J. Oosterlaan

## Abstract

Neurocognitive functioning is crucial for child development, but the relevance of interplay between neurocognitive functions in childhood remains poorly understood. This explores the relevance of neurocognitive network organization in childhood by the application of network theory to neurocognitive data at the individual level. A community sample of children (N = 132), between 6 and 18 years of age (M = 11.4) performed computerized neurocognitive testing. Neurocognitive connectivity was calculated between each pair of test scores, creating individual connectivity matrices after which graph theory was applied to determine global network organization and local network organization. The relation between demographics and neurocognitive network organization was investigated with correlation analysis. The relevance for intelligence (Wechsler short forms) and behavioral problems (Strength and Difficulties Questionnaire) was assessed with multivariate regression. K-means clustering was applied to investigate the correspondence between typical neurocognitive network organizations and conventionally assessed neurocognitive performance. The results show that global as well as local neurocognitive network organization is related to age, intelligence and behavior problems. Moreover, children from neurocognitive clusters with typical configurations of global network organization also differed in terms of conventionally assessed neurocognitive performance. In conclusion, this study provides cross-sectional evidence suggesting the presence of developmental reorganization of the interplay between neurocognitive functions. Neurocognitive network organization is also related to crucial aspects of functioning in children (intelligence, behavior problems) and the level of conventionally assed neurocognitive performance. Thereby, this study shows that individual network analysis provides a complementary view on the child functioning and may hold relevance for a better understanding of child development and the influence of neuropathology on daily life functioning.

## Introduction

Neurocognitive functioning is a crucial aspect of child development, importantly contributing to everyday activities,^1^ behavioral and social functioning,^2^ academic achievement^3^ and later socio-economic functioning.^4^ A considerable body of research has investigated neurocognitive functioning of healthy and diseased children, which has shaped our understanding of normal and compromised child development.^5, 6^ Nevertheless, the available research has neglected the complex interplay between neurocognitive functions, thereby providing a limited view on neurocognitive functioning and development in children.

Neurocognitive functions are known to heavily interact in order to facilitate behavior^7^. Nevertheless, studies have long disregarded neurocognitive functioning as a complex network of interconnected functions, until the influence of network theory changed the dominant paradigm of neuroscience from a localization approach to a network approach.^8^ Network theory is a powerful method to study the organization of comprehensive and complexly interconnected structures or processes and has been widely adopted in neuroscience to study the organization of structural and functional brain networks.^9^ In recent years, network theory has also been applied to study the organization of the neurocognitive network, unlocking a novel dimension in our understanding of neurocognitive functioning. Indeed, recent studies have presented evidence suggesting that neurocognitive network organization is sensitive for the impact of neurological disorders in children and older adults.^10–13^

Previous studies have applied network theory to investigate neurocognitive network organization in children with epilepsy. These studies provided evidence indicating aberrant neurocognitive network organization in children with epilepsy compared to healthy peers. More specifically, the neurocognitive network of children with epilepsy showed a lower degree of clustering, indicating reduced interplay between neurocognitive functions. Likewise, other studies have found evidence supporting neurocognitive reorganization effects in older adults with mild cognitive impairment and Alzheimer disease.^13^ More specifically, patients with pathological aging show weaker specialization in the neurocognitive network. Taken together, these studies underline the relevance of neurocognitive network organization for our understanding of disease. Nevertheless, the existing studies applied network theory to neurocognitive data at the group level, using correlations between variables *across* individuals as a measure of connectivity. Although this approach can importantly contribute to our understanding of neurocognitive network organization at group level, implementing network theory to individual neurocognitive assessment data, would enable the possibility to determine the neurocognitive network organization at the individual level, to investigate the relevance of inter-individual differences and would open up the possibility to evaluate neurocognitive network organization for clinical purposes.

In previous work,^14^ we showed that neurocognitive network organization can be determined at the individual level (coined as the ‘neurocognome’), with considerable agreement with the previously deployed group-based approach in healthy young adults. The validity of the individual network approach was also supported by considerable overlap in the neurocognitive network across random subgroups of the study sample, indicating relative stability of the neurocognitive network between individuals, as well as considerable inter-individual differences in the organization of the network. Moreover, parameters of neurocognitive network organization showed relevance for other important domains of functioning, such as intelligence and behavioral functioning. Interestingly, neurocognitive network organization accounted better for the prevalence of behavior problems in healthy young adults than did conventional measures of neurocognitive functioning. Although network analysis may unlock a new dimension in our understanding of child functioning, the neurocognitive network has to date not been studied in children at individual level.

The current study aims to explore the relevance of the neurocognome in a community sample of children between school age and late adolescence. We will investigate the relation between demographic characteristics and neurocognitive network organization in order to determine whether neurocognitive network organization has potential to deepen our knowledge of child development, sex differences and socio-economic status (SES). We will also investigate the relevance of neurocognitive network organization for other crucial aspects of child functioning, such as intelligence and behavioral functioning. Lastly, in order to improve our understanding of the relation between neurocognitive network organization and the conventional view on neurocognitive functioning, we will explore the existence of neurocognitive clusters of children with specific configuration of neurocognitive network organization, and compare these groups of children in terms of their conventional neurocognitive performance. The results of the present study will reveal the potential of the neurocognome to deepen our understanding of pediatric neurocognitive functioning.

## Methods

### Participants

This observational study used a diverse community sample of children recruited between July 2021 and May 2023 through various sources including primary schools, secondary schools, sports organizations and social organizations throughout the Netherlands, and through a network of trusted partners collaborating with Amsterdam University Medical Centers. Inclusion criteria were: (1) aged between 6 and 18 years; (2) native control over the Dutch language; and (3) inhabitant of the Netherlands. Exclusion criteria were: (1) inability to understand test instructions; (2) severe primary or secondary sensory or motor impairment interfering with neurocognitive testing. We strived to include at 20 participants for each 2-year age band in the age range between 6-18 years, totaling to a minimum total sample size of (6 * 20 =) 120 subjects.

### Procedure

Interested organizations circulated an invitation to participate in our study. Children and parents of children < 16 years of age with interest in the study were informed by a member of the research team and were provided an information letter. After oral consent, either a visit to the Emma Children’s Hospital was planned, or a visit to our mobile laboratory (‘Emma Brain Bus’) was planned. At time of the visit and before research procedures were initiated, written informed consent was collected from children aged >11 years and parents of children aged <16 years. Ethical approval was given by the medical ethical committee of the Amsterdam University Medical Centers (location AMC, NL76915.018.21) and the trial was registered at the International Clinical Trials Registry Platform (NL9574). The data corresponding to this manuscript (doi: 10.17026/dans-z5w-q4st) is published online at https://dans.knaw.nl/nl/data-stations/life-health-and-medical-sciences/. This manuscript follows STROBE reporting guidelines for observational studies.^15^

#### 1.1 Measures

##### Demographic information

Demographic information (i.e. age, sex, socio-economic status; SES) and medical background (i.e. self-reported clinical diagnoses) was collected using a custom online questionnaire. SES was defined as the mean highest educational level of parents based on a 7-point scale ranging from no education (1) to postdoctoral education (8).

##### Intelligence & behavioral functioning

Intelligenceandbehavioraflunctioningweremeasuredtocapturecruciadl omainsof functioning with relevance to neurocognitive functioning. Intelligence was measured by a short form of the Wechsler Intelligence Scale for Children-V^17^ for child younger than 16 years of age or the Wechsler Adult Intelligence Scale-IV^18^ for children as from 16 years of age, involving the Vocabulary, Similarities, Block Design and Matrix Reasoning subtests. Full-scale IQ (FSIQ) estimated with this short form has excellent validity (*r*s ≥ .82) and reliability (*r* ≥. 92).^19^ Behavioral functioning was measured using the Dutch version of the Strength and Difficulties Questionnaire (SDQ), a widely used tool allowing the measurement of behavioral and emotional problems and more specifically, the presence of internalizing problems (emotional and peer relationship problems; Internalizing scale) and externalizing problems (conduct and attention problems; Externalizing scale). The parent version of the SDQ was completed by parents of all children younger than 16 years of age. The SDQ is known to have adequate validity, internal consistency and inter-rater reliability.^20^

##### Neurocognitive functioning

Neurocognitive functioning was measured using the ‘Emma Toolbox for Neurocognitive Functioning’, an in-house developed composition of computerized tests based on well-established neuroscientific paradigms with proven validity and reliability. The Emma Toolbox objectively assesses information processing and attention, learning and memory, executive functions and visuomotor integration. The battery is designed to maximize the use of built-in experimental task manipulations (to isolate specific neurocognitive functions) and parametric difficulty manipulation (to assess the influence of increasing task load on performance).

Furthermore, the test battery is optimized for network analysis by a symmetrical design in the verbal and visual domain (i.e. identical test design for tests assessing functions in the verbal and visual domain), minimizing the influence of test design on task performance. See the Supplementary Information for detailed descriptions of the neurocognitive tests used, and see Table 1 for an overview of the variables that were extracted from the data. The Emma Toolbox is programmed in Python using OpenSesame Software^21^.

**Table 1.**
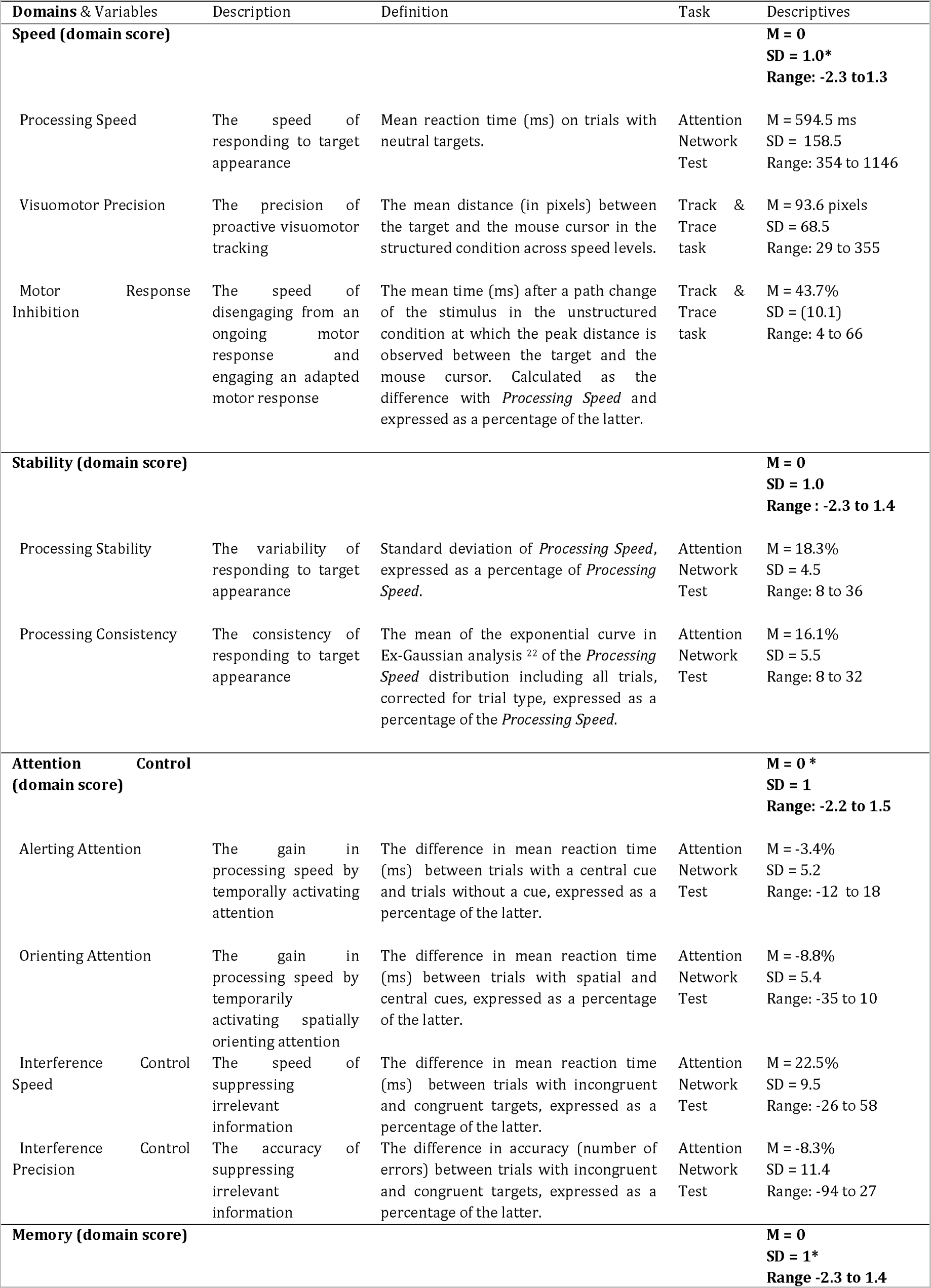
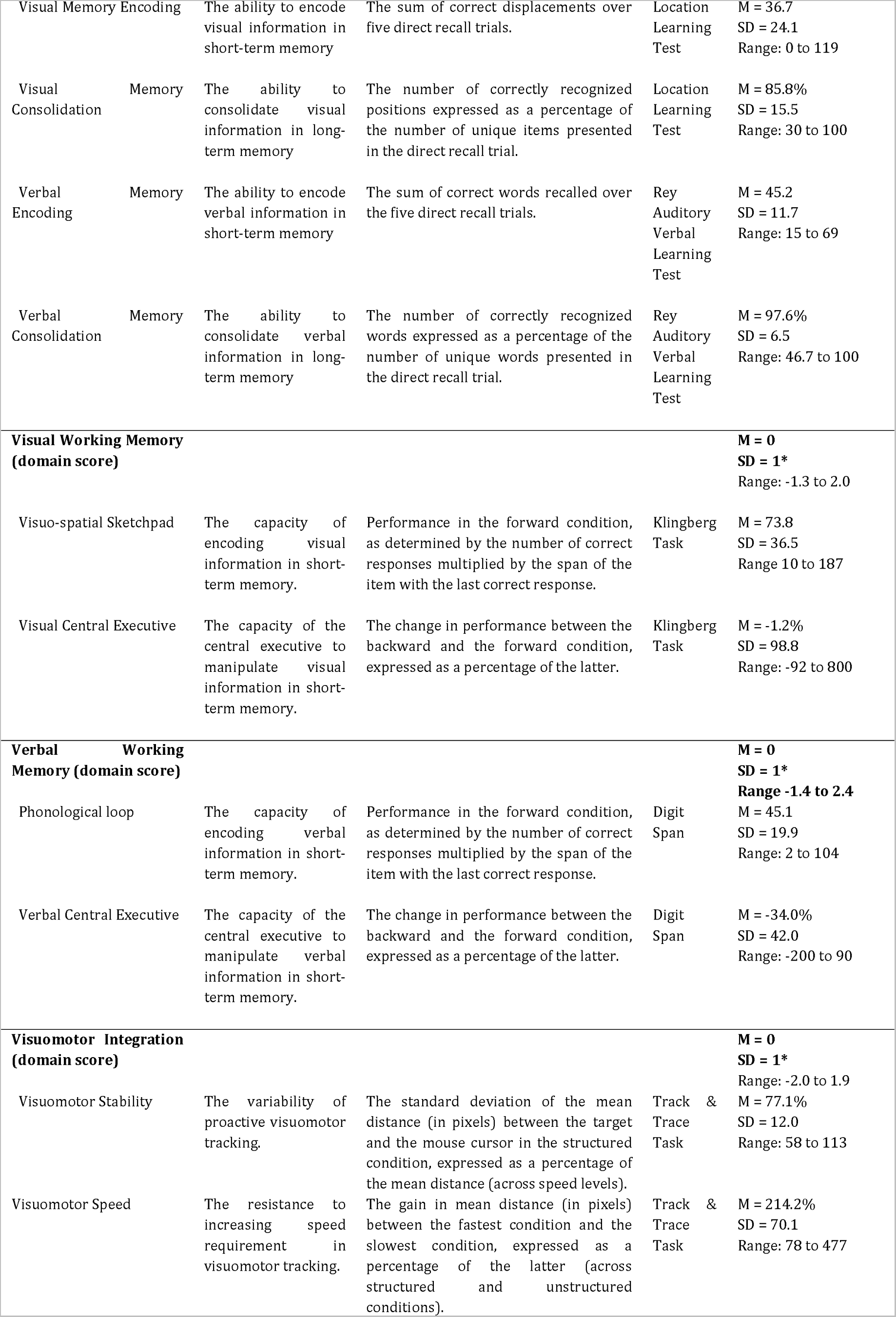
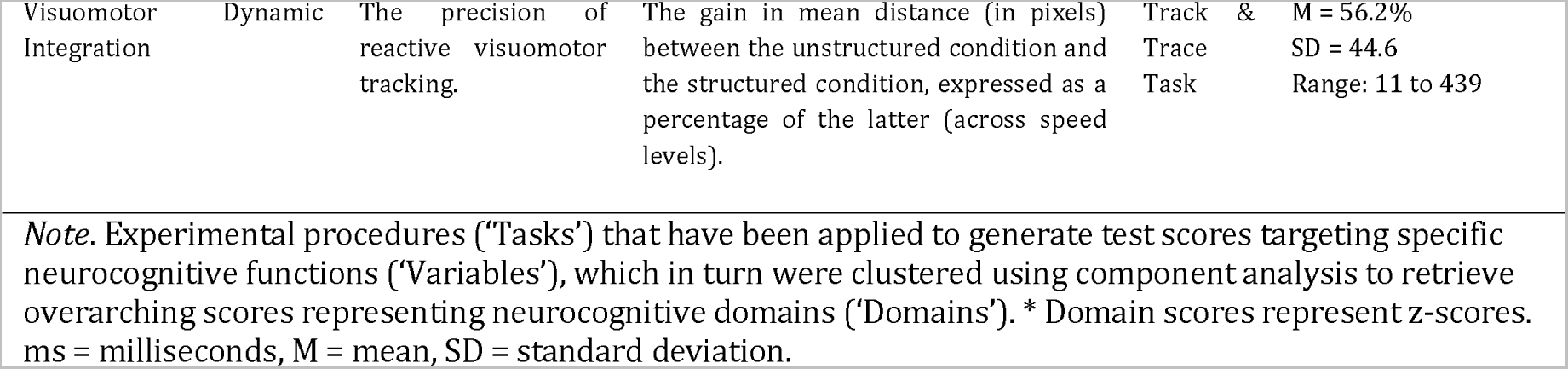
Overview of the neurocognitive domains, variables, definitions and tasks used for network construction

##### Pre-processing

The neurocognitive test data were subjected to a pre-processing pipeline to prepare the data for network analysis in R^23^. First, all neurocognitive test scores were normalized, to allow comparison of test scores on the same scale (i.e. z-scores). Second, if applicable, the directionality of the neurocognitive test scores was adapted with higher values corresponding to better task performance. Third, domain scores were extracted from the data using principal component analysis with varimax rotation from the ‘Psych’ package. ^24^Thenumberof components (i.e. neurocognitive domains) to extract was determined using the elbow method and the cumulative variance explained from the eigenvalues histogram (*R*^2^ > 70%). Subsequently, each component was labeled as a neurocognitive domain based on the set of test scores that made the strongest contribution to the components, as measured in terms of *n*^2^. The boundary for this set of test scores was set at the largest drop in *n*^2^ between two subsequent variables in the scree plot. For each component, the unweighted mean of this set of test scores was used to calculate neurocognitive domains scores, which are displayed in Table 1.

##### Connectivity matrices

The pre-processed neurocognitive data (the domain scores supplemented with the neurocognitive test scores) were then subjected to a processing pipeline to create individual connectivity matrices using a procedure that is described in detail in a previous study. In summary, the following steps were undertaken. The first step was to calculate neurocognitive connectivity between all neurocognitive test scores described in Table 1. Based on the idea that highly connected functions are likely to function at a comparable level, neurocognitive connectivity is defined by the absolute difference in corresponding test scores on a standardized scale (z-scores). This method was used to construct a weighted neurocognitive connectivity matrix for each participant. The second step was to reduce the influence of chance connectivity. By the definition of our neurocognitive connectivity measure, the resulting matrices may be sensitive to ‘chance connectivity’, meaning that two neurocognitive test scores have comparable z-scores by chance, instead of an underlying connection between the corresponding functions. As a solution, we use group-level correlations to limit the analysis to connections that are robustly identified at the group-level. As a third step, we further reduced the influence of redundant connections by using a range of thresholds to select the strongest connections in each individual connectivity matrix. To account for the influence of arbitrary threshold selection on network organization (Qi & Meesters, (2015), network parameters (see below) were calculated at the individual level across a range of matrix thresholds from the 75% to the 25% strongest connections, with steps of 5%. Further analyses were performed in every connectivity matrix at each threshold level. As a consequence of the thresholding procedure, neurocognitive functions could in theory be ‘disconnected’ (i.e. isolated from the network). As the fourth and last step, we reconnected isolated neurocognitive functions to the backbone of the network, determined by the minimum spanning tree.^25^

#### 1.2 Network organization

Graph theory was applied to the resulting individual weighted connectivity matrices to study the organization of the neurocognitive network at the individual level (i.e. neurocognome). Graph theory is an influential method in connectivity analysis that describes the organization of a network according to the distribution of links (i.e. connections) between nodes (i.e. in this case neurocognitive functions), see a review for more background on graph theory. ^26^Global network parameters describe the organization of the network as a whole, while local network parameters describe characteristics of a specific node in the network. All network parameters were calculated using the ‘igraph’ and ‘qgraph’ packages in R.^27, 28^

##### Global network organization

The following network parameters were used to assess global network organization: *strength, modularity, assortativity, characteristic path length, transitivity, and smallworldness. Strength* describes the sum of connectivity values in the network, there by reflecting the average coherence in the network. *Modularity* describes the subdivision of the network into modules, which are groups of relatively strong interconnected nodes, assessing the level of specialization in the network. *Assortativity* describes the tendency of nodes with a high number of links (‘hubs’) to connect to other hubs, reflecting hierarchy in the network. *Characteristic path length* describes the average number of links between each node in the network and any other node in the network, reflecting integration in the network. *Transitivity* describes the ratio between triangles (three nodes connected in a closed triangle) to triplets (three nodes connected in an open triangle), reflecting clustering in the network. *Smallworldness* describes the ratio between clustering and integration in the network, relative to a random network. *Smallworldness* is known to be an important feature of network efficiency.^29^ Smallworld networks are significantly more clustered than random networks while having comparable integration, reflected by a *smallworldness* value greater than one.^27^

##### Local network organization

The *hubness score* was used as a measure of local network organization, reflecting the relative importance of nodes (i.e. neurocognitive functions) in the network as measured by centrality (i.e. authority) in the network. The hub score was calculated according to the Kleinberg algorithm.^30^

### Statistical analysis

The statistical analysis was performed in R. ^23^The influence of outliers was reduced by winsorizing with the ‘DescTools’ package and data missing at random (≤3.8%) were imputed using the ‘Mice’ package with classification and regression trees. The individual connectivity matrices were averaged and the resulting connectivity matrix was visualized as a neurocognitive network using the ‘qgraph’ package.^31^ We ran a set of validation procedures to determine (1) the value of the connectivity measures, (2) the correspondence of our individual network approach with a group-based approach and (3) the stability of the neurocognitive network across random groups of individuals. Full details on these analyses are provided in the Supplementary information. Furthermore, the relation between network threshold and global network parameters was investigated by repeated measure ANOVA using network threshold as a within-subject factor. Lastly, neurocognitive network hubs were determined as the 25% neurocognitive functions with the highest hubness score across network thresholds, as calculated using the area under the curve.

Three sets of variables were used to assess aspects of neurocognitive functioning: (i) conventional neurocognitive measures (z-score for each neurocognitive domain, *k* = 7, also see Table 1) measuring performance on neurocognitive tests; (ii) global network parameters (*strength, modularity, assortativity, characteristic path length, transitivity,*and *smallworldness, k* = 6) measuring global network organization; and (iii) local network organization (hubness score for each neurocognitive domain, *k* = 7) measuring relative importance of each neurocognitive domain in the network. For global and local network parameters we used the area under the curve to obtain a single value for each parameter across network thresholds.

In order to investigate the relevance of neurocognitive network organization for developmental effects, sex differences and socio-economic status, Pearson correlations were calculated between demographic characteristics (age, sex and SES), conventional neurocognitive measures, global network parameters and local network parameters.

The relevance for intelligence and behavior and emotional problems was assessed using multiple linear regression models in the ‘caret’ package.^32^ FSIQ, the SDQ Internalizing problems score and the SDQ Externalizing problems score served as dependent variables. We selected four sets of predictors: conventional measures, global network parameters, local network parameters and all predictor sets combined. For each set of predictors, we ran a separate regression model, with demographic predictors (age, sex and SES) added to the model. In order to prevent overfitting, pre-selection of predictors was performed using recursive feature selection. Subsequently, cross-validated stepwise feature selection in backward direction was performed based on Akaike’s Information Criterion, selecting the best performing model using five-fold cross-validation with each ten repeats. The number of predictors in the model was maximized at 10 observations/predictor.^33^ Model performance was assessed by the explained variance (adjusted *R*^2^). To statistically compare model performance between predictor sets, 95%-confidence intervals (CIs) were calculated around each estimate of model performance using bootstrapping (based on 5000 samples), where the model based on conventional neurocognitive measures served as reference model.

Lastly, we attempted to increase our understanding of the relation between neurocognitive network organization and conventionally assessed neurocognitive performance by investigating whether children with differential network organization would also differ on conventionally assessed neurocognitive performance. Therefore, we used k-means cluster analysis on global network parameters to determine subgroups of children with a similar configuration of global network characteristics. (i.e. neurocognitive clusters). Children from each neurocognitive clusterwerecomparedtotheotherchildrenonglobalnetworkparametersinorderto determine the identity the of the subgroups in term of neurocognitive network organization. Subsequently, the subgroups were also compared on local network parameters and conventional measures in order to determine how global neurocognitive organization clusters translate into differences in local network organization and conventionally assessed neurocognitive performance.

Statistical analyses were two-sided with alpha = .05. Multiple comparisons were accounted for in correlation analyses and group comparisons by FDR-correction at the level of conventional measures, global network parameters and local network parameters.

## Results

### Study sample

Among the study sample of children (*n* = 132), the majority had the Dutch nationality (76.5%), the average age was 11.6 years (range: 6.0-18.9), sexes were approximately evenly distributed (43% female) and the educational level was on average 6.3 points on a 1-8 scale (range: 4.5-8.0) indicating that the parents of participants had on average received higher education. With regard to self-reported diagnoses, *n* = 3 had a learning disorder (dyslexia: *n* = 3,), *n* = 6 had a behavioral disorder (ADHD: *n* = 3, anxiety disorder: *n* = 1, autism: *n* = 2), while *n* = 1 of the participants had a neurological disorder (suspected myoclonic epilepsy of infancy: n = 1). The study sample had intelligence in the higher average range (FSIQ: M = 110.9, SD = 12.5) and prevalence of behavior problems (SDQ total score: M = 6.1, SD = 4.9) corresponded well with Dutch normative values (6-11 years: M = 8.2, SD = 6,2; 12-18 years: M = 6.6, SD = 5,3).^34^

### Neurocognitive network organization

The average neurocognitive network as reconstructed at the individual level is displayed in Figure 2. The planned validation procedures are reported in the Supplemental Information. The results confirm the expectation that neurocognitive connectivity is higher between neurocognitive functions that are part of the same neurocognitive domain as compared to neurocognitive functions that are part of different neurocognitive domains (*t*[131] = 7.2, *p* < .001). This finding supports the validity of the used definition of neurocognitive connectivity. The validity of the individual network approach is further supported by significant overlap of the individual neurocognitive networks with the group-based network approach (61.9% overlap, 95%-CI: 54.1 - 69.6). Lastly, we found evidence for stability of the individual neurocognitive networks between randomly selected subgroups of our study sample (64.3% overlap, 95%-CI: 52.8 - 75.8). Taken together, these findings suggest that the measure of neurocognitive connectivity behaves as expected and as previously observed in young healthy adults^14^, giving rise to individual neurocognitive networks that have considerable consistence across existing approaches as well as across individuals.

**Figure 1.**
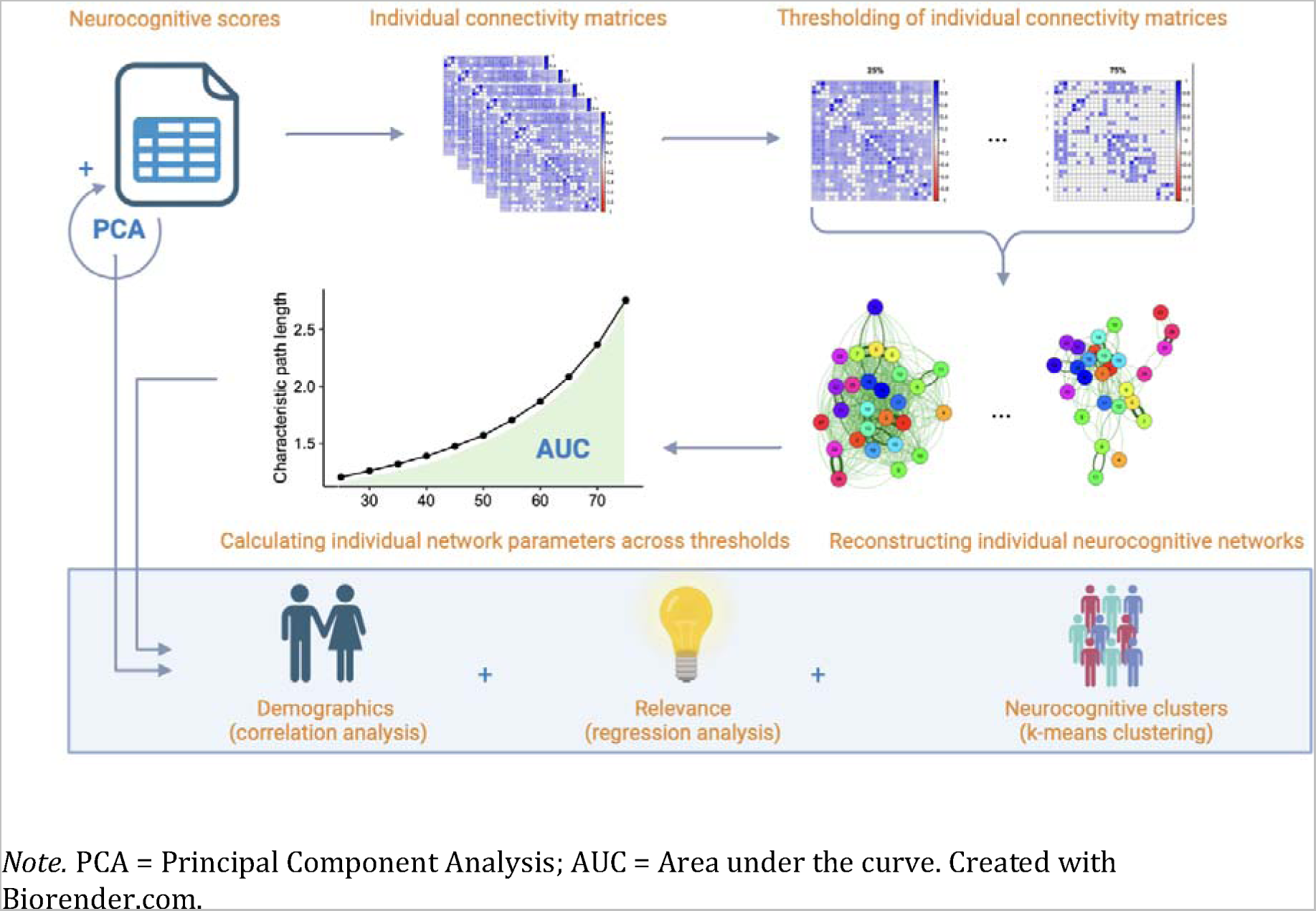
Overview of data processing and analyses.

**Figure 2.**
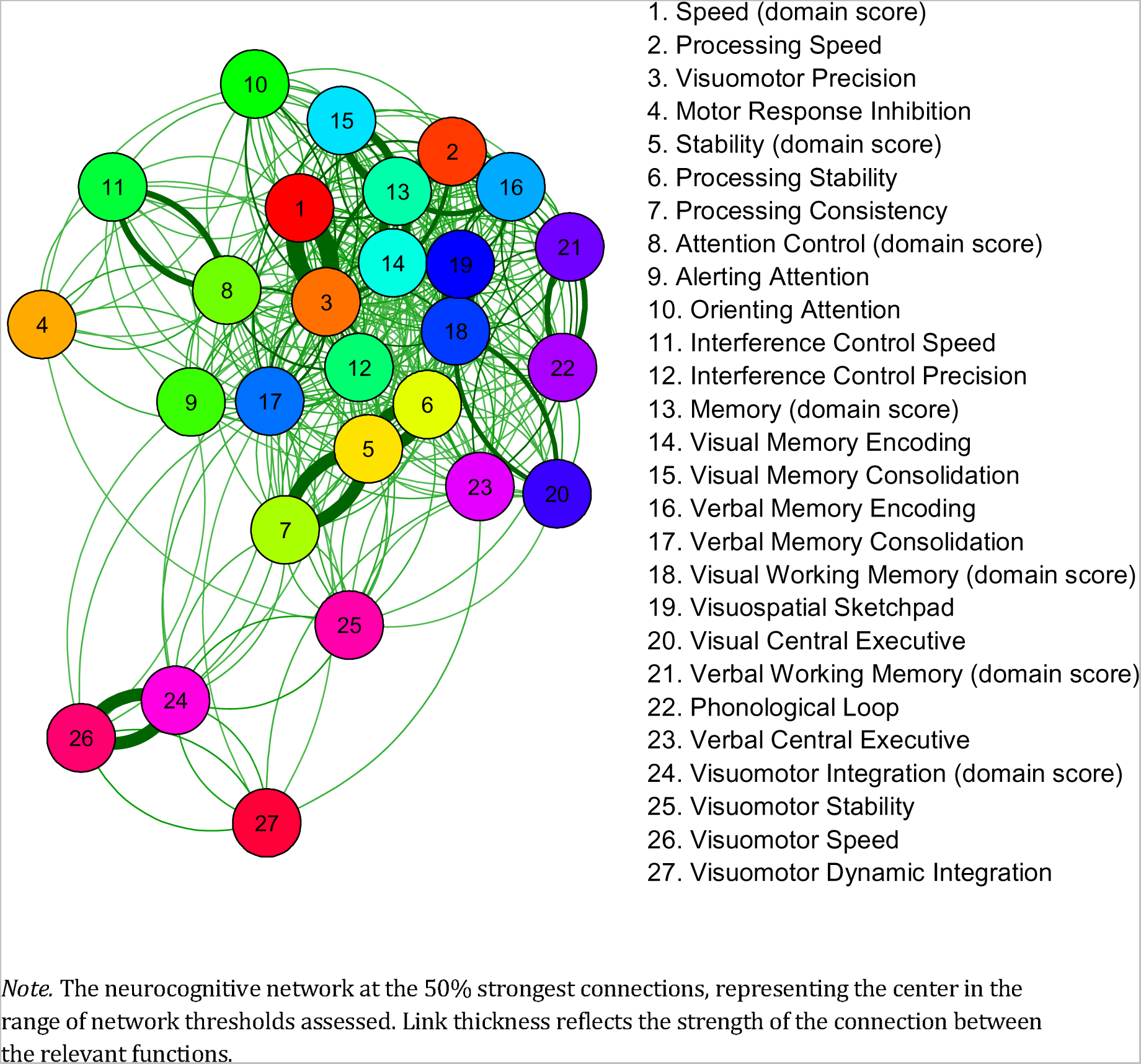
The average neurocognitive network as reconstructed at the individual level.

#### Global network organization

Global neurocognitive network organization was determined across network thresholds (Figure S2), revealing that more sparse networks have: lower network *strength* (indicating lower coherence in the network); higher modularity (indicating higher specialization); higher *assortativity* (indicating more prominent hierarchy); longer *characteristic path length* (indicating lower integration); lower *transitivity* (indicating lower level of clustering); higher *smallworldness* (indicating that sparser networks tend to favor clustering over integration). These findings replicate earlier findings in healthy young adults^14^.

#### Local network organization

Figure 3 displays the importance of neurocognitive functions in the network as measured by the hubness score (area under the curve across network thresholds). *Visuomotor Precision* had the highest hubness score, indicating that this process has the most central position in the neurocognitive network. The top 25% hubs in the network further included *Speed* (domain score), *Memory* (domain score), *Visual Memory Encoding, Visual Working Memory* (domain score), *Processing Speed, Visuospatial Sketchpad* .

**Figure 3.**
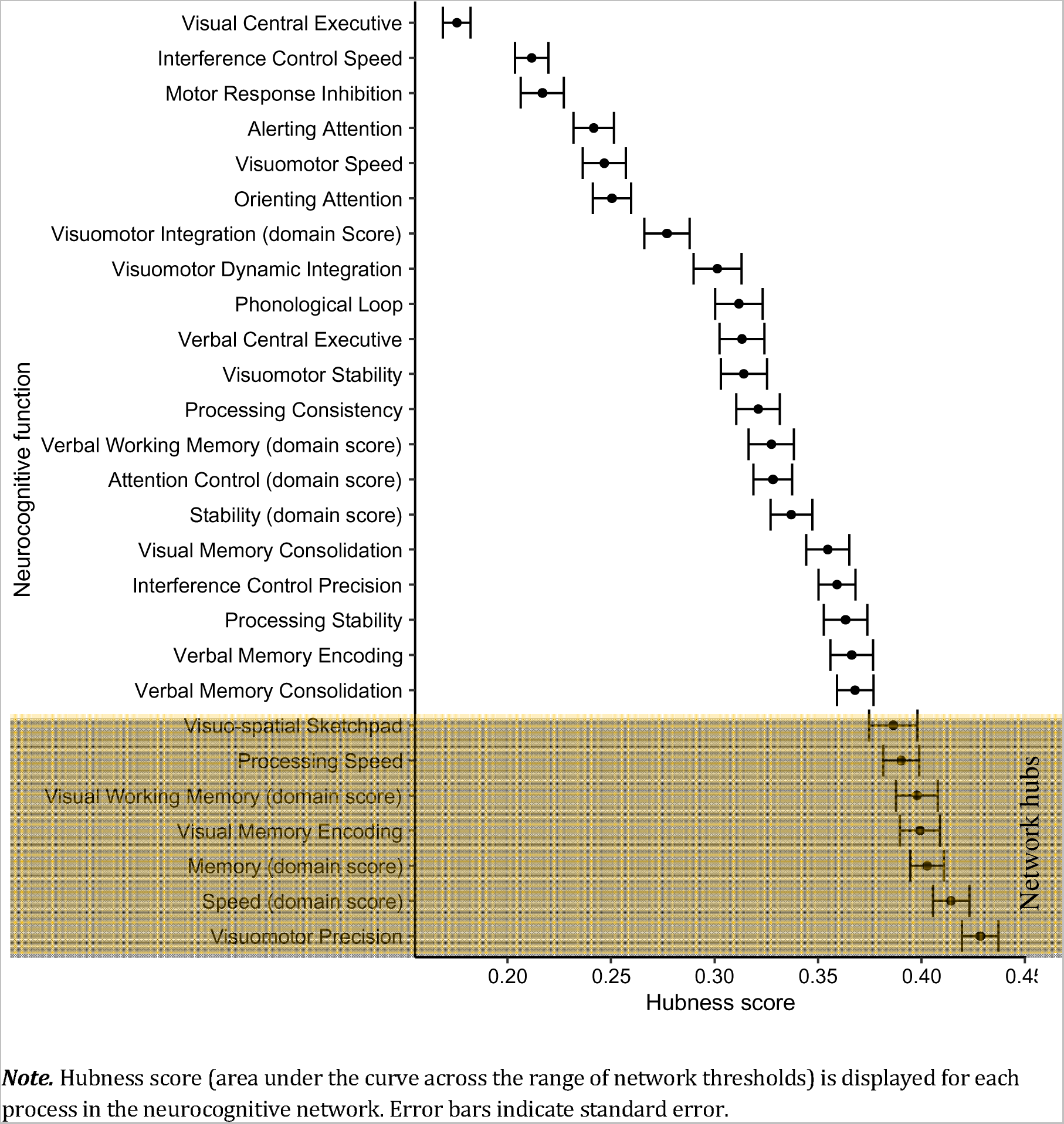
Local network organization over the range of network thresholds.

### Relation to demographics

Among conventional neurocognitive measures (Figure 4), higher age was significantly associated with higher scores for all neurocognitive domains (*rs*: .39-.67, *p*s < .001), with the exception of the Attention Control domain (*r* = .14, *p* = .10) and the Visuomotor Integration domain (*r* = −.16, *p* = .08). Regarding global network parameters (Figure 5), higher age was related to lower *strength* (*r* = −.26, *p* = .01), indicating that older children have lower coherence among the neurocognitive test scores. No significant relations were found between age and *characteristic path length* (*r* = .02, *p* = .80), *transitivity*, (*r* = .10, *p* = .50), *modularity* (r = −0.04, p = .74), *assortativity (r* = .19 *, p* = .08 ) or *smallworldness* (*r* = −.06, *p* = .72), suggesting that integration, clustering, specialization, hierarchy and smallworld organization of the neurocognitivenetworkarerelativelystableacrosschildhood.Intermsoflocalnetwork organization (Figure 6), we found that higher age was associated with higher hubness of Speed domain scores (*r* = .36, *p* < .001) and lower hubness of scores on the Visual Working Memory domain (*r* = −0.30, *p* < .001) and the Verbal Working Memory domain (*r* = −.21, *p* = .04). These findings indicate that in older children, speed plays a more central role in the neurocognitive network, whereas visual and verbal working memory play a less central role in the network.

**Figure 4.**
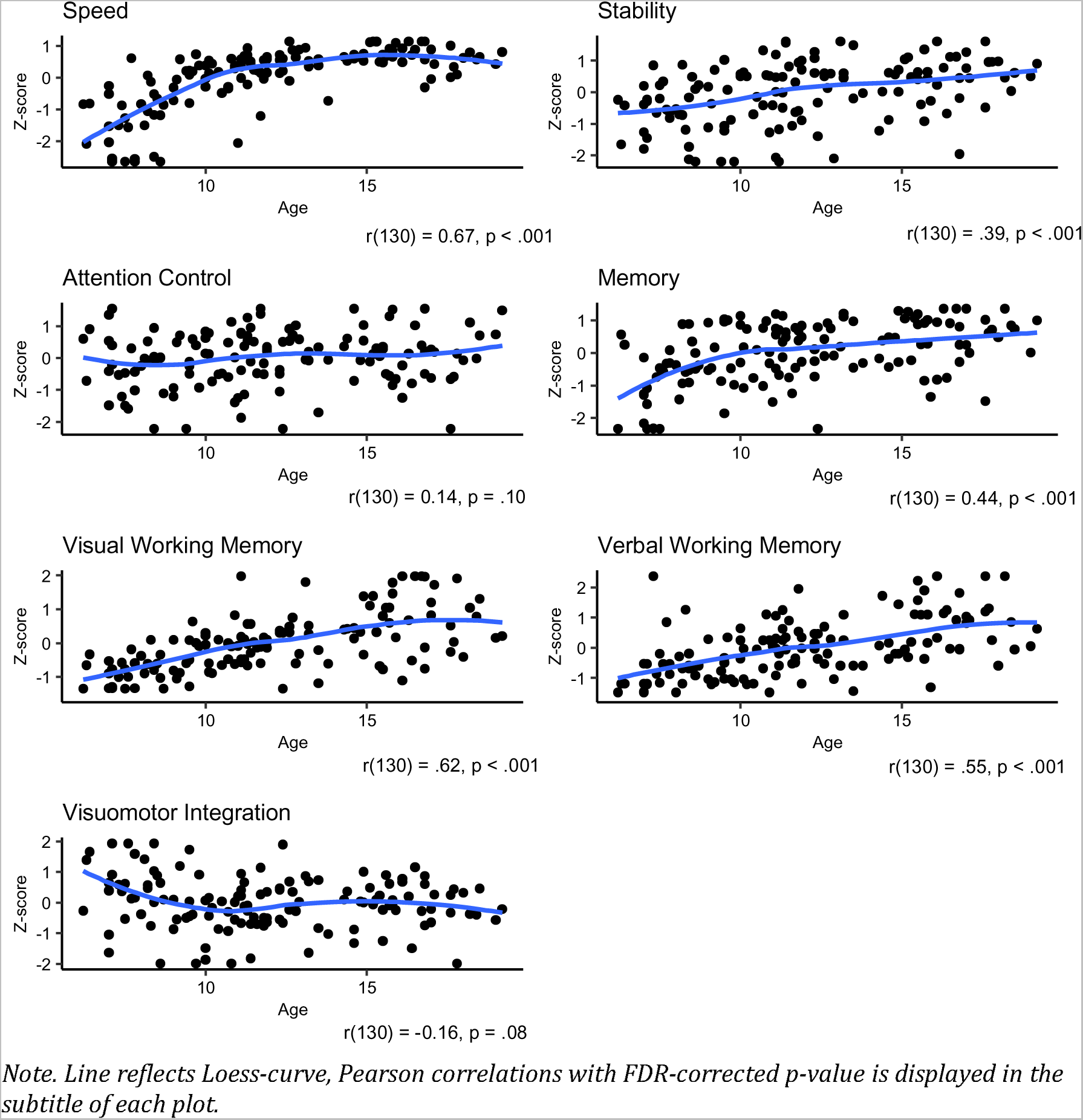
Age and conventional neurocognitive domain measures.

**Figure 5.**
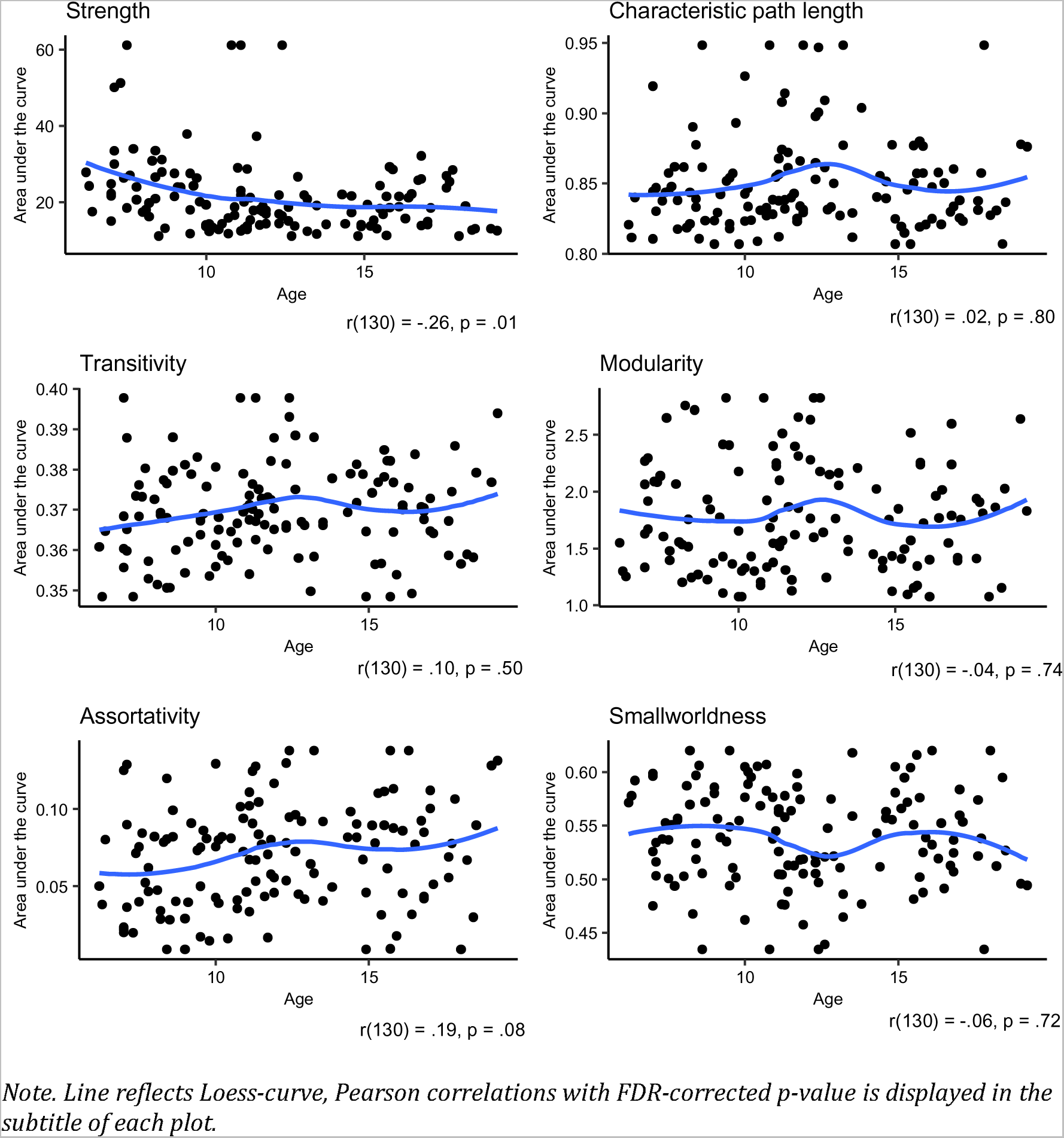
Age and global network organization.

**Figure 6.**
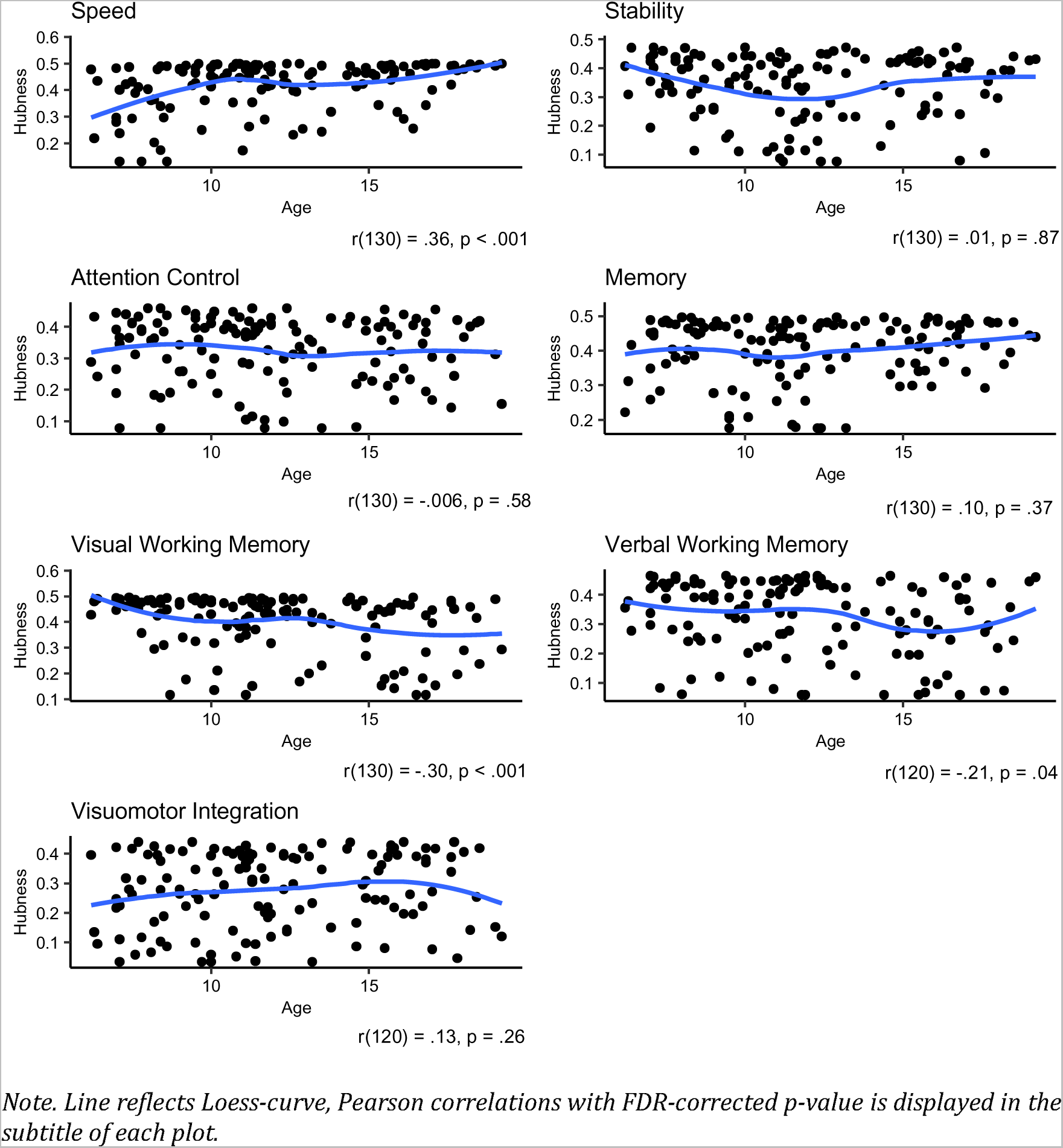
Age and local network organization.

Regarding sex (Figures S6-S8), we found no significant difference between children with female sex and children with male sex on the conventional neurocognitive measures (−0.3 ≤ *t*s ≤ 1.5, *p*s > .12), global network parameters (−0.3 ≤ *t*s ≤ 2.2, *p*s ≥ .17) or local network parameters (−0.8 ≤ *t*s ≤ 0.7, *p*s ≥ .90), with exception of children with female sex having higher hubness of scores on the visuomotor integration domain than children with male sex (*t* = 3.1, *p* = .01). This suggests that boys and girls do not differ on conventional neurocognitive performance and global network organization, while in term of local network organization, visuomotor integration may play a more central role in the neurocognitive network of girls as compared to boys.

Regarding SES (Figures S9-S11), we found no significant associations with conventional neurocognitive measures (−.03 ≤ *r*s ≤ .21, *p*s > .06), global network parameters (−.15 ≤ *r*s ≤ .18, *p*s > .26), and local network parameters (−.12 ≤ *r*s ≤ .19, *p*s > .23), with the exception of higher SES being associated with higher conventional scores for attention control (*r* = .25, *p* = .02) memory performance (*r* = .03, *p* = .007). These results suggest that SES is not related to global network organization or local network organization, yet that children with higher SES have higher conventional performance for attention control and memory functions.

### Relevance of neurocognitive network organization

The relevance of conventional neurocognitive measures (z-scores) and neurocognitive network organization was assessed by regression analysis with intelligence, and internalizing and externalizing problems acting as outcome variables. Demographic variables (age, sex, and SES) were added as covariates in all models.

#### Intelligence

The reference model based on conventional neurocognitive measures (z-scores) was predictive of intelligence (Table 2, *R*^2^ = 32.5%), where higher z-scores on the Memory domain were related to higher FSIQ. Global network organization also had predictive value for intelligence (*R*^2^ = 27.8%). Lower *modularity* and lower *strength* was related to higher FSIQ, suggesting that less pronounces specialization and coherence in the neurocognitive network are related to higher intelligence. None of the local network parameters was included as a significant predictor of intelligence.

**Table 2.** Relations between neurocognitive parameters and intelligence.

| Predictor set Model | Predictors | Statistics |  | Performance |  | Fit AIC |
| --- | --- | --- | --- | --- | --- | --- |
|  |  | B (SE) | P | Adj. R<sup>2</sup> | 95%-CI |  |
| Conventional measures | **Memory (z-score)** | **.46 (.07)** | **<.001** | 32.5% | Reference | 331.7 |
|  | **Age** | **-.27 (.08)** | **.001** |  |  |  |
|  | **SES** | **.26 (.09)** | **.001** |  |  |  |
| Global network parameters | **SES** | **.40 (.09)** | **<.001** | 27.8% | 17.3 – 40.3% | 344.7 |
|  | **Modularity** | **-.29 (.01)** | **.04** |  |  |  |
|  | **Strength** | **-.24 (.09)** | **.009** |  |  |  |
|  | Age | -.15 (.08) | .15 |  |  |  |
| Local network parameters | **SES** | **.43 (.08)** | **<.001** | 18.7% | 11.1 – 32.2% | 354.3 |
|  | Visual Working Memory (hubness) | -.12 (.08) | .15 |  |  |  |
| All | **Memory (z-score)** | **.50 (.07)** | **<.001** | 42.3% | 31.2 – 53.8% | 295.4 |
|  | **Age** | **-.39 (.10)** | **<.001** |  |  |  |
|  | **Modularity** | **-.35 (.12)** | **.005** |  |  |  |
|  | **Smallworldness** | **-.32 (.16)** | **.04** |  |  |  |
|  | Characteristic path length | .29 (.14) | .05 |  |  |  |
|  | **SES** | **-.24 (.07)** | **.002** |  |  |  |
|  | Transitivity | .18 (.14) | .12 |  |  |  |
|  | **Visumotor Integration (hubness)** | **.15 (.07)** | **.04** |  |  |  |
|  | Verbal Working Memory (z-score) | .15 (.09) | .10 |  |  |  |
|  | Speed (hubness) | -.12 (.09) | .13 |  |  |  |

**Note.** $n = 132$ . SE = standard error; CI = confidence interval, AIC = Aikake's Information Criterion; SES = socio-economic status.

When combining parameters sets, the resulting hybrid model was predictive of intelligence (42.5%) and involved significant predictors assessing conventional neurocognitive performance (z-score of the Memory domain) as well as global network organization (*modularity* and *smallworldness*). Based on the 95% confidence intervals around model performance, the model for conventional neurocognitive measures had better performance as compared to the model with local network parameters. Likewise, the model with combined parameter sets had better performance than the models with global and local network parameters.

#### Internalizing behavior problems

Conventional measures of neurocognitive functioning were not captured as significant predictors in the model for internalizing problems (Table 3, *R*^2^ = 3.4%) and neither were global network parameters (*R*^2^ = 3.4%). Local network parameters did show some predictive value for internalizing problems (*R*^2^=7.6%).More specifically, higher hubness of the scores on the Visuomotor Integration domain was related to more internalizing problems, indicating that children in whom visuomotor integration functions have a more central position in the network, tended to have more internalizing problems.

**Table 3.** Relations between neurocognitive parameters and internalizing behavior problems.

| Predictor set | Model | Statistics |  | Performance |  | Fit AIC |
| --- | --- | --- | --- | --- | --- | --- |
|  |  | B (SE) | P | Adjusted R<sup>2</sup> | 95%-CI |  |
| Conventional measures | SES | -.18 (.09) | .05 | 3.4 | Reference | 313.4 |
| Global network parameters | SES | -.18 (.09) | .05 | 3.4 | 0.1 – 16.8 | 313.4 |
| Local network parameters | **Visuomotor Integration (hubness)** | **.20 (.09)** | **.03** | 7.6 | 1.3 – 18.9 | 310.5 |
|  | Attention Control (hubness) | -.18 (.09) | .05 |  |  |  |
| All | **Visuomotor Integration (hubness)** | **.20 (.09)** | **.03** | 7.6 | 1.3 – 18.9 | 310.5 |
|  | Attention Control (hubness) | -.18 (.09) | .05 |  |  |  |

**Note.** Subsample of children younger than 16 years of age (n = 110). SE = standard error; CI = confidence interval, AIC = Aikake's Information Criterion; SES = socio-economic status.

When combining parameter sets, the resulting model accounted for internalizing problems (*R*^2^ = 7.6%) and included only one significant predictor assessing local network organization (hubness of Visuomotor Integration). This finding suggests that local network organization relatively accounts best for the prevalence of internalizing problems, although no differences in performance were observed between the models assessed.

#### 3.5.3 Externalizing behavior problems

The model for conventional measures of neurocognitive functioning (Table 4, *R*^2^ = 17.2%) captured only z-scores for the Memory domain as a significant predictor. Children with poorer conventional performance on the Memory domain tended to have more externalizing problems. Global network organization was related to externalizing problems as well (*R*^2^ = 16.6%), where higher *transitivity* was related to more externalizing problems. This suggests that children with stronger clustering in the neurocognitive network tended to have more externalizing problems. Local network parameters were also related to externalizing problems (*R*^2^ = 17.0%), with hubness of the Verbal Working Memory domains as significant predictor. Higher hubness of the Verbal Working Memory domain was related to more externalizing problems. This finding suggests that children with more externalizing problems also had a more central role for verbal working memory performance.

**Table 4.** Relations between neurocognitive parameters and externalizing behavior problems.

| Predictor set | Model | Statistics |  | Performance |  | Fit<br>AIC |
| --- | --- | --- | --- | --- | --- | --- |
|  |  | B (SE) | P | Adjusted<br>R<sup>2</sup> | 95%-CI |  |
| Conventional measures | **Memory (z-score)** | **-.23 (.09)** | **.02** | 17.2 | Reference | 302.4 |
|  | Sex (Female) | -.16 (.09) | .09 |  |  |  |
|  | Attention Control (z-score) | -.15 (.09) | .13 |  |  |  |
| Global network parameters | **SES** | **-.19 (.09)** | **.02** | 16.6 | 4.0 – 34.4 | 301.2 |
|  | **Transitivity** | **.18 (.09)** | **.04** |  |  |  |
|  | Sex (Female) | -.17 (.09) | .07 |  |  |  |
|  | Strength | .17 (.09) | .06 |  |  |  |
| Local network parameters | **Verbal Working Memory (hubness)** | **.22 (.09)** | **.02** | 17.0 | 5.0 – 32.9 | 302.7 |
|  | **Sex (Female)** | **-.22 (.09)** | **.02** |  |  |  |
|  | SES | -.17 (.09) | .07 |  |  |  |
|  | Attention Control (hubness) | -.16 (.09) | .09 |  |  |  |
| All | **Memory (z-score)** | **-.21 (.08)** | **.01** | 24.7 | 9.8 – 38.5 | 294.0 |
|  | **Transitivity** | **.21 (.08)** | **.02** |  |  |  |
|  | **Attention Control (z-score)** | **-.18 (.09)** | **.04** |  |  |  |
|  | Attention Control (hubness) | -.16 (.09) | .06 |  |  |  |
|  | Verbal Working Memory (hubness) | .14 (.09) | .13 |  |  |  |

**Note.** Subsample of children younger than 16 years of age (n = 110). SE = standard error; CI = confidence interval, AIC = Aikake's Information Criterion; SES = socio-economic status.

The hybrid model combining all parameters (*R*^2^ = 24.7%) captured significant predictors assessing conventional neurocognitive performance (z-score for the Memory and Attention Control domains) and global network organization (*transitivity*). This suggests that global network measures (i.e. transitivity) add to the relevance of conventional neurocognitive measures for the understanding of externalizing problems in children, although performance of the combined model did not significantly differ from the model using only global or local network parameters.

### Neurocognitive clusters

In order to investigate the relation between neurocognitive network organization and inter-individual differences in terms of conventional neurocognitive performance, we explored the existence of clusters of children with a comparable configuration of global network measures using k-means cluster analysis. Clusters of children with comparable network configuration were first compared on global network measures in order to determine the identity of the neurocognitive clusters (Figure 7). Subsequently the neurocognitive clusters were compared on local network parameters (Figure 8) and conventional neurocognitive measures (Figure 9) in order to determine how different configurations of global network organization translate into local network organization and conventional neurocognitive performance. The clustering algorithm provided evidence for the identification of three neurocognitive clusters.

**Figure 7.**
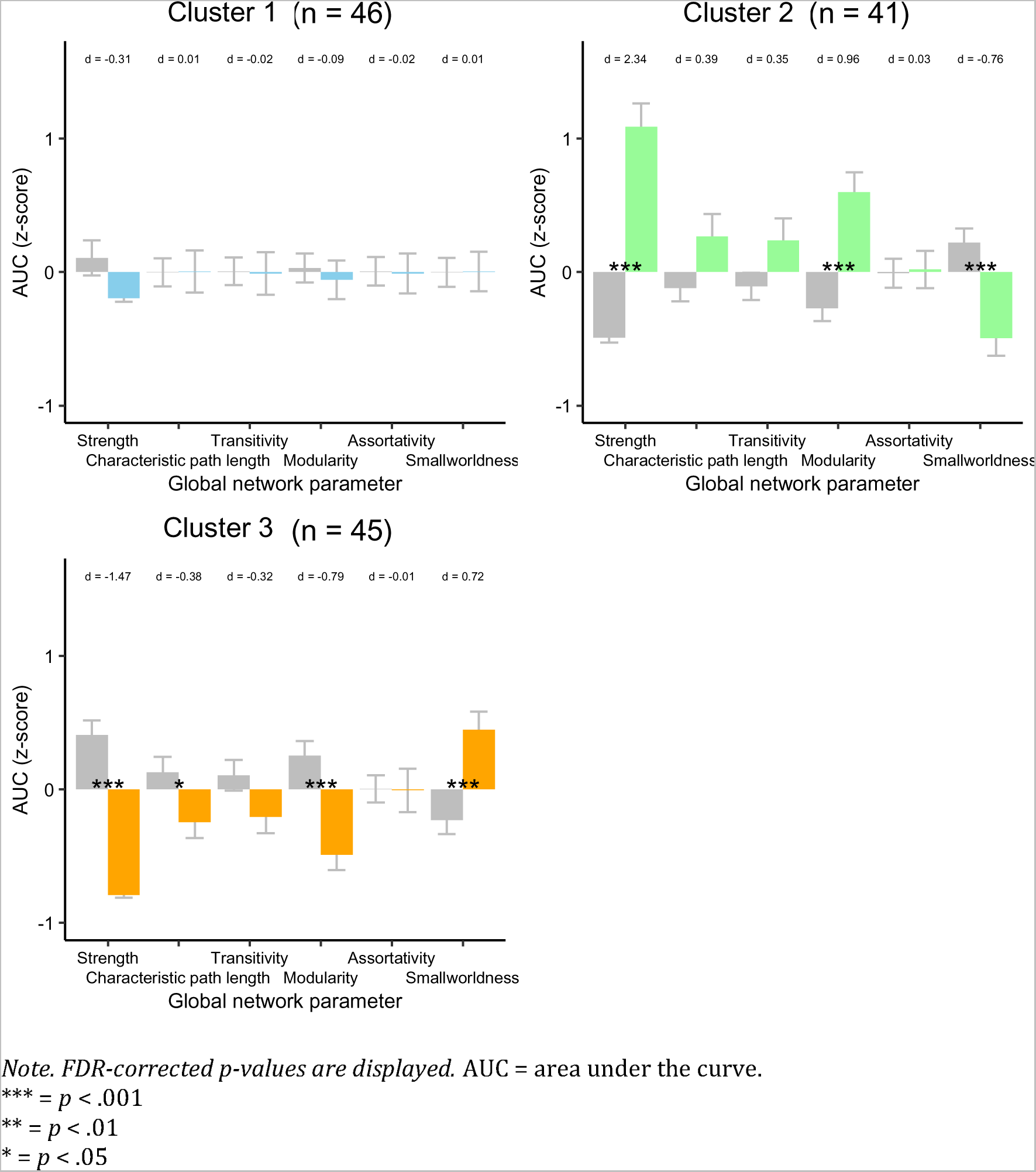
Neurocognitive clusters and global network organization

**Figure 8.**
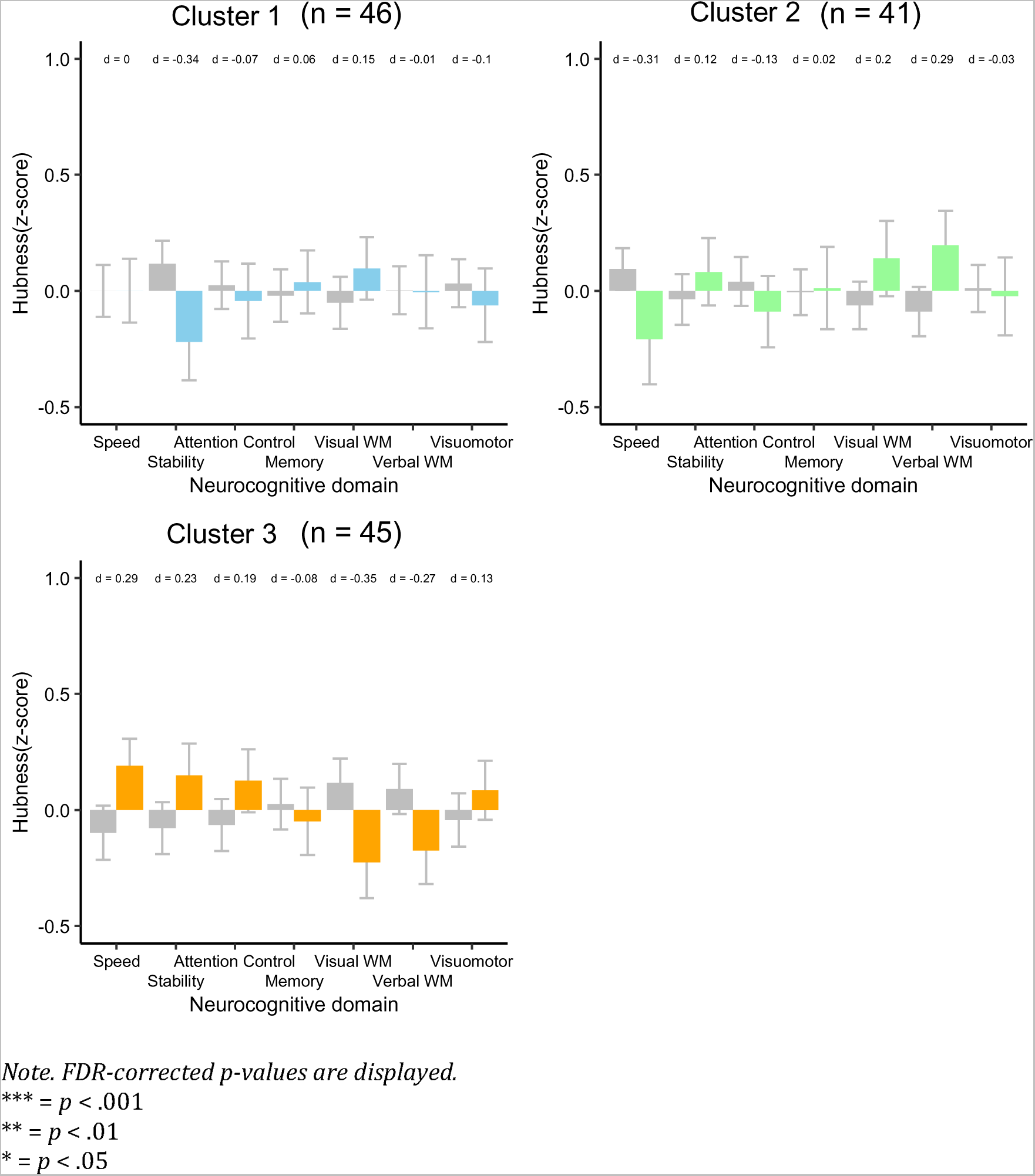
Neurocognitive clusters and local network organization

**Figure 9.**
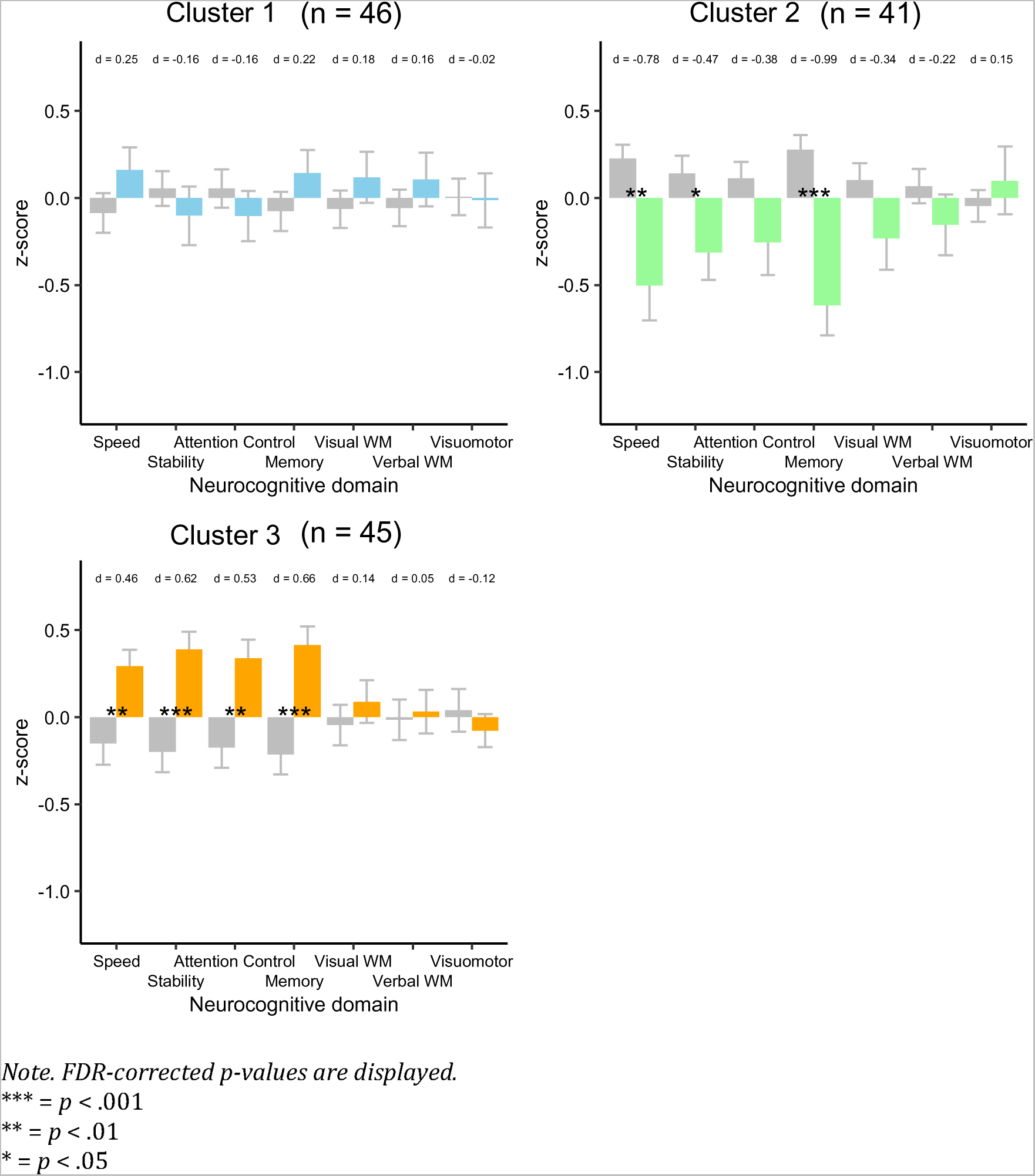
Neurocognitive clusters and conventional neurocognitive performance.

#### Global network organization

Compared to the other children, children in neurocognitive cluster 1 (*n* = 46, 35%) did not differ on any of the global network organization measures (*p*s > .05, −0.02 < *d*s < 0.31). In contrast, children in neurocognitive cluster 2 (n = 41, 31%) had higher *strength* (*p* < .001, *d* = 2.34), higher *modularity* (*p* < .001, *d* = 0.96), lower *smallworldness* (*p* < .001, *d* = −0.76) than the other children. Vice versa, children in neurocognitive cluster 3 (*n* = 44, 34%) had lower *strength* (*p* < .001, *d* = −1.47), lower *modularity,* (*p* < .001, *d* = −0.79), lower characteristic path length (*p* < .05, *d* = −0.38) and higher *smallworldness* (*p* < .001, *d* = 0.72). These findings suggest that clusters of children exist with a certain configuration of global network organization, primarily characterized by differences in coherence when considering effect sizes of group differences. One cluster is characterized by average global network organization (cluster 1), while the other groups are characterized by opposing configurations regarding coherence in the network (cluster 2: high, cluster 3: low), specialization (cluster 2: high, cluster 3: low) and smallworldness (cluster 2:low, cluster 3: high).

#### Local network organization

There were no differences in local network organization (Figure 8) between children from each neurocognitive cluster and the other children (*p*s > .05, −0.34 ≥ *d*s ≥ 0.39). Taken together, these findings suggest that differential configurations of global network organization do not translate into the relative importance of specific neurocognitive functions in the neurocognitive network.

#### Conventional neurocognitive measures

Compared to all other children, children in neurocognitive cluster 1 did not differ from the other children on any of the domain scores (*p*s > .05, −0.16 ≥ *d*s ≥ 0.24). Children in neurocognitive cluster 2 had lower z-scores (Figure 9) on the domains Memory (*p* < .001, *d* = −0.99), Speed (*p* < .01, *d* = −0.78) and Stability (*p* < .05, *d* = −0.47). Children from neurocognitive cluster 3 had higher z-scores than the other children on domains Memory (*p* < .001, *d* = 0.66), Stability (*p* < .01, *d* = 0.62), Attention Control (*p* < .05, *d* = 0.53) and Speed (*p* < .01, *d* = 0.46). Taken together, these findings indicate that differential configurations of global network organization among children translate into differential performance in terms of conventionally assessed neurocognitive functioning.

## Discussion

This study investigated the value of neurocognitive network organization in childhood by application of network theory to neurocognitive data, exposing ‘the neurocognome’ at the individual level. The findings of this study reveal that neurocognitive network organization is related to age between middle and late childhood, suggesting developmental reorganization of interplay between neurocognitive functions. Neurocognitive network organization was further found to be relevant for daily life functioning, more specifically in terms of intelligence and behavioral functioning. Lastly,theresultsofthisstudyprovideinsightintherelationship between network organization and conventional measures of neurocognitive functioning, by showing that children with diverging configurations of global network organization, also differ on conventional measures of neurocognitive functioning. Taken together, the findings indicate that neurocognitive network organization may provide a complementary view on neurocognitive functioning in childhood, by providing insight into the relatively unexplored aspect of interplay between neurocognitive functions.

This study contributes to an emerging field of study^35^, providing evidence reflecting the value of neurocognitive network organization for daily life functioning in young adults, ^14^understanding of the impact of epilepsy in childhood,^10–12^ the influence of aging and dementia in late adulthood,^13, 36, 37^ and neuropharmacological effects in children with autism spectrum disorder.^38^ The current study extends the existing literature by deploying neurocognitive network analysis at the individual level, enabling the exploration of inter-individual differences in neurocognitive network organization to provide deeper understanding of neurocognitive functioning in a community sample of children.

Analyses aimed at the relation between demographics and neurocognitive network organization indicate minor relevance of sex and socio-economic status. In contrast, we found that age was related to neurocognitive network organization. Regarding global network organization, older children had lower coherence (*integration)* in the neurocognitive network. This cross-sectional finding may suggest that maturation of neurocognitive functioning as from middle childhood is characterized by proliferation of specific strengths in the neurocognitive profile, rather than a general improvement in neurocognitive performance. These findings represent novel evidence from network theory to support the cognitive differentiation hypothesis.^39^ In terms of local network organization, the results indicate that the connectivity of speed-related functions with other functions (*hubness*) increases with older age, while the connectivity of visual and verbal working memory functions decreases. Taken together, these cross-sectional findings are suggestive of developmental reorganization of the neurocognitive network.

This study further showed that global as well as local neurocognitive network organization have relevance for daily life functioning in terms of intelligence and behavior problems. Global network organization was related to intelligence, with lower coherence (*strength)* and lower specialization (*modularity*) in the neurocognitive network being related to higher intelligence. Although speculative, this result might suggest that a more mature configuration of the neurocognitive network (i.e. lower *strength*) also sets the stage for optimized performance. Local network organization was modestly related to internalizing problems, where greater connectivity of visuomotor functions with other functions (*hubness*) in the network was associated with higher prevalence of internalizing problems. Interestingly, the relation between visuomotor functioning and internalizing problems does not manifest when using conventional measures of performance (r = .07, p = .48), suggesting that the relative importance of visuomotor functions in the neurocognitive network has more relevance to internalizing problems that the conventional neurocognitive performance for these functions. Global as well as local network organization were related to externalizing problems. More specifically, more externalizing problems were accounted for by stronger clustering (*transitivity)* in the neurocognitive network, suggesting that a more fragmented neurocognitive network organization characterized by global hyperconnectivity among closely related neurocognitive functions and by local hyperconnectivity of verbal working memory functions may contribute to emergence of externalizing problems. It may be speculated that this global hyperconnectivity reflects a reduced integration in the network, with relative disconnection of higher-order regulatory functions (i.e. attention control functions), which may then increase the likelihood of behaviors that are disruptive to the social environment.

In order to gain more insight in the relation between neurocognitive network organization and conventional measures of neurocognitive performance, we investigated how differential configurations of global network organization translate into local network organization and conventionally assessed neurocognitive performance. A data-driven clustering algorithm revealed three clusters of children with differential global network organization. The pattern of results suggest that configurations of global network organization is independent of local network organization, suggesting that the connectivity of specific neurocognitive functions in the network does not dictate the organization of the network as a whole and vice versa. In contrast, we found that differential configurations of global network organization are related to conventionally assessed measures of neurocognitive performance. More specifically, children with stronger coherence (*strength*), stronger specialization (*modularity*) and lower smallworldness (cluster 2), had poorer conventionally assessed neurocognitive performance (i.e. speed, stability and memory functions). Children with average coherence, specialization and smallworldness (cluster 1) also had average conventionally assessed neurocognitive performance. Children with weaker coherence, weaker specialization, greater smallworldness and lower integration (*characteristic path length*, cluster 3) had better conventionally assessed neurocognitive performance (i.e. speed, stability, attention control and memory functions). Taken together, these findings indicate that certain configurations of global network organization are associated with better conventionally assessed neurocognitive performance, and suggest that lower coherence in the neurocognitive network, in combination with a lower degree of specialization and greater smallworldness, provides an optimized neurocognitive network organization for the facilitation of neurocognitive performance as measured in a conventional way.

This study has strengths and weaknesses. First, we collected a considerable community sample of children between middle and late childhood. Moreover, we used an innovative method to apply network theory on neurocognitive data at the individual level. It should be noted that our method uses an indirect behavioral measure of neurocognitive connectivity, meaning that under some circumstances the assumption of connectivity may be violated (e.g. two measurements have similar z-scores by chance, instead of reflecting an underlying connection). Nevertheless, this also accounts for more established magnetic resonance imaging based measures of connectivity used to reconstruct structural brain networks and functional brain networks^40^. Another point of attention is that the reconstruction of the neurocognitive network may be dependent on the composition of tests that produce the neurocognitive data. We argue to have used a balanced battery of neurocognitive tests, covering the major neurocognitive domains with symmetrical test designs for verbal and visual assessments. Nevertheless, it remains unknown to what extent the findings of this study generalize to neurocognitive networks as reconstructed using other neurocognitive datasets. The study results do support the validity and relevance of the current approach to model the neurocognitive network.

In conclusion, this study provides cross-sectional evidence suggesting the presence of developmental reorganization of the interplay between neurocognitive functions. Neurocognitive network organization is also related to crucial aspects of functioning in children (intelligence, behavior problems) and optimized (conventional) neurocognitive performance. Multiple lines of evidence from this study point to the importance of coherence (*strength*) in the child’s neurocognitive network, reflecting more dominant proliferation of specific strengths (and weaknesses) in the neurocognitive profile. The hypotheses regarding neurocognitive functioning and development in children raised by this study await replication in longitudinal studies. Nevertheless, the findings from this study indicate that individual neurocognitive network analysis provides a complementary view on child functioning, may hold relevance for a better understanding of typical child development as well as the influence of neuropathological impacts on child functioning.

## Data Availability

The data corresponding to this manuscript (doi: 10.17026/dans-z5w-q4st) is published online at https://dans.knaw.nl/nl/data-stations/life-health-and-medical-sciences/.

## Supplementary information

**Table S1.** Description of the neurocognitive tests.

| Task | Description |
| --- | --- |
| Track & Trace task<sup>41</sup> | A moving target stimulus is presented on the computer screen. Subjects are instructed to keep the mouse cursor on the center of the target in a structured condition (predictable, circular path) and in an unstructured condition (unpredictable, random path) at four linearly increasing target speeds. Cursor position is measured using a gaming mouse (1000 Hz refresh rate) and the speed of the moving stimulus is corrected for the system refreshing rate. |
| Attention Network Test<sup>42</sup> | Target stimuli pointing left or right are presented on a computer screen. Subjects are instructed to respond as quickly as possible to the direction of a target stimulus by pressing the corresponding button. Performance is influenced by the presentation of cues (central, spatial) and manipulation of target flanker congruency (neutral, congruent, incongruent). The measurement of reaction times is corrected for system latency. |
| Location Learning Test<sup>43</sup> | A grid (5 x 50) with 10 semi-randomly placed visual stimuli (the template) is presented five times. The subject has to reproduce the template by replacing each visual stimulus in an empty grid, directly after each presentation (direct recall) and after a 30 minutes interval (delayed recall). Lastly, the subject has to recognize the correct location of each visual stimulus in the grid, among 10 distractor locations (recognition). |
| Rey Auditory Verbal Learning Test (Dutch Version)<sup>44</sup> | A list of 15 words is auditory presented five times. The subject has to reproduce as many words as possible directly after each presentation (direct recall) and after an interval of 30 minutes (delayed recall). Lastly the subject has to select the presented words among 15 distractors (recognition). |
| Klingberg task<sup>45</sup> | A sequence of stimuli is presented on a four by four digital grid. Subjects are required to repeat the sequence in the order of presentation (forward) or reversed order (backward) by clicking on the relevant locations in the grid. The difficulty increases every other trial, by increasing the length of the sequence or increasing the difficulty of the virtual trajectory of the yellow dots. Performance in each condition is defined by the span (the difficulty level of the last correct trial) multiplied by the total number of correct trials. |
| Digit Span<sup>46</sup> | Subjects are required to repeat a sequence of numbers presented auditory in the order of presentation (forward condition) or reversed order (backward condition). The difficulty increases every other trial, by increasing the length of the sequence of digits. Performance in each condition is defined by the span (the difficulty level of the last correct trial) multiplied by the total number of correct trials. |

**Note.**

### Value of the connectivity measure

We first tested the expectation that neurocognitive connectivity should be higher for connections between neurocognitive variables that are more strongly related to each other. In line with the expectation, the results (Figure S2) show that average neurocognitive connectivity was higher for connections within neurocognitive domains as compared to connections across neurocognitive domains (t(131) =7.2, p < .001). This finding suggests that our measure of connectivity (i.e. intra-individual differences in z-scores) may be a useful (indirect) measure of neurocognitive connectivity.

**Figure S1.**
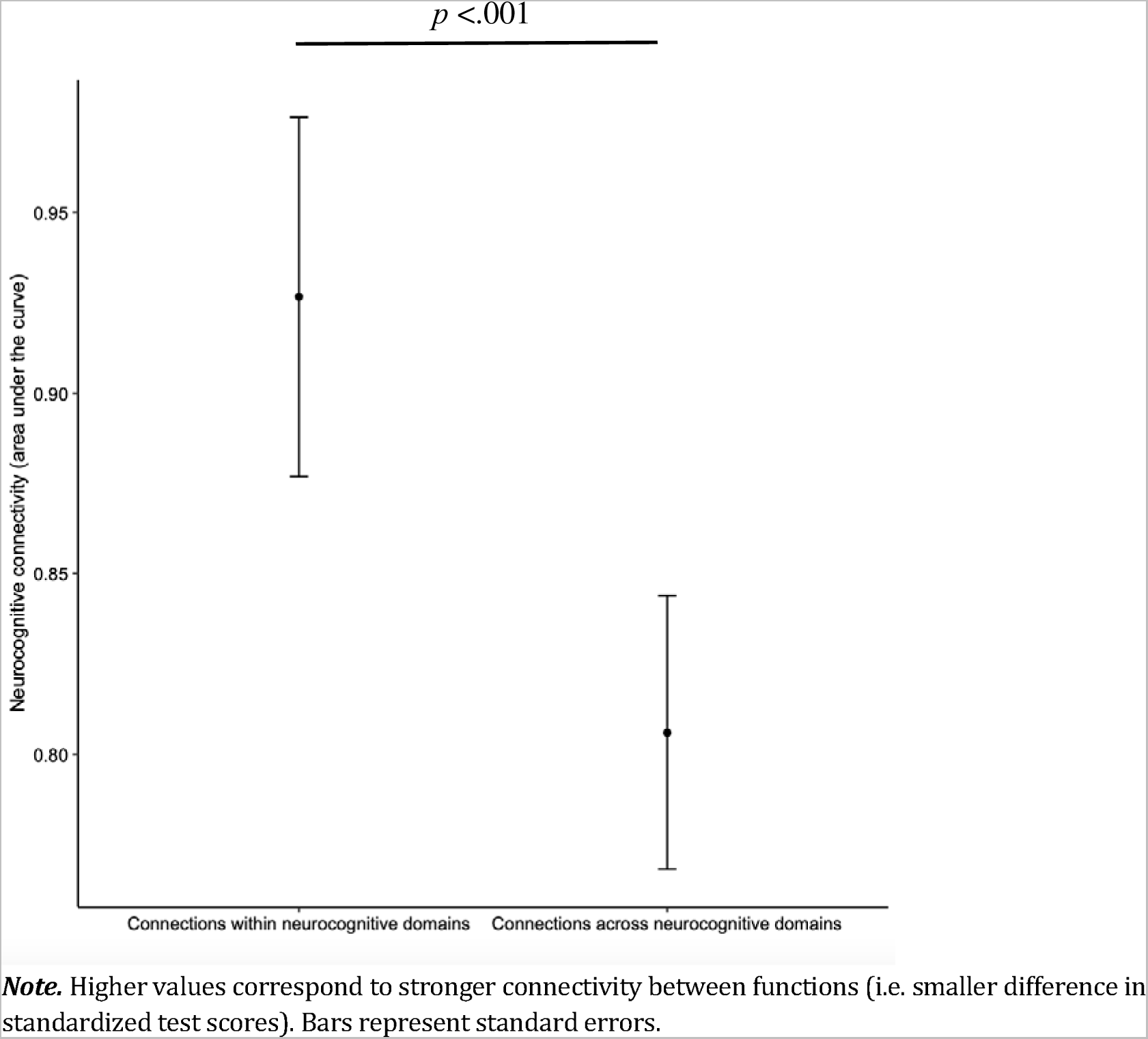
Neurocognitive connectivity of connections within vs. across neurocognitive domains.

### Validity of the individual network approach

We investigated the validity of our individual network approach as compared to the previously used group-based network approach. Therefore, we reconstructed the neurocognitive network using both methods and determined their correspondence by the overlap between the two networks (Figure S3). The results show that the networks have considerable overlap, sharing 61.9% of their connections (bootstrap 95%-CI: 54.1% −69.7%). The observed overlap was found to significantly exceed the overlap that would be expected on chance level (6.25%). This finding indicates that the individual network approach produces a neurocognitive network that strongly corresponds to a network that would be retrieved when using group-based Pearson correlations as a measure of connectivity, supporting the validity of our individual network approach. At the same time, the differences between the networks created by both approaches are also considerable, suggesting that the individual network approach also captures unique variation in the neurocognitive network.

**Figure S2.**
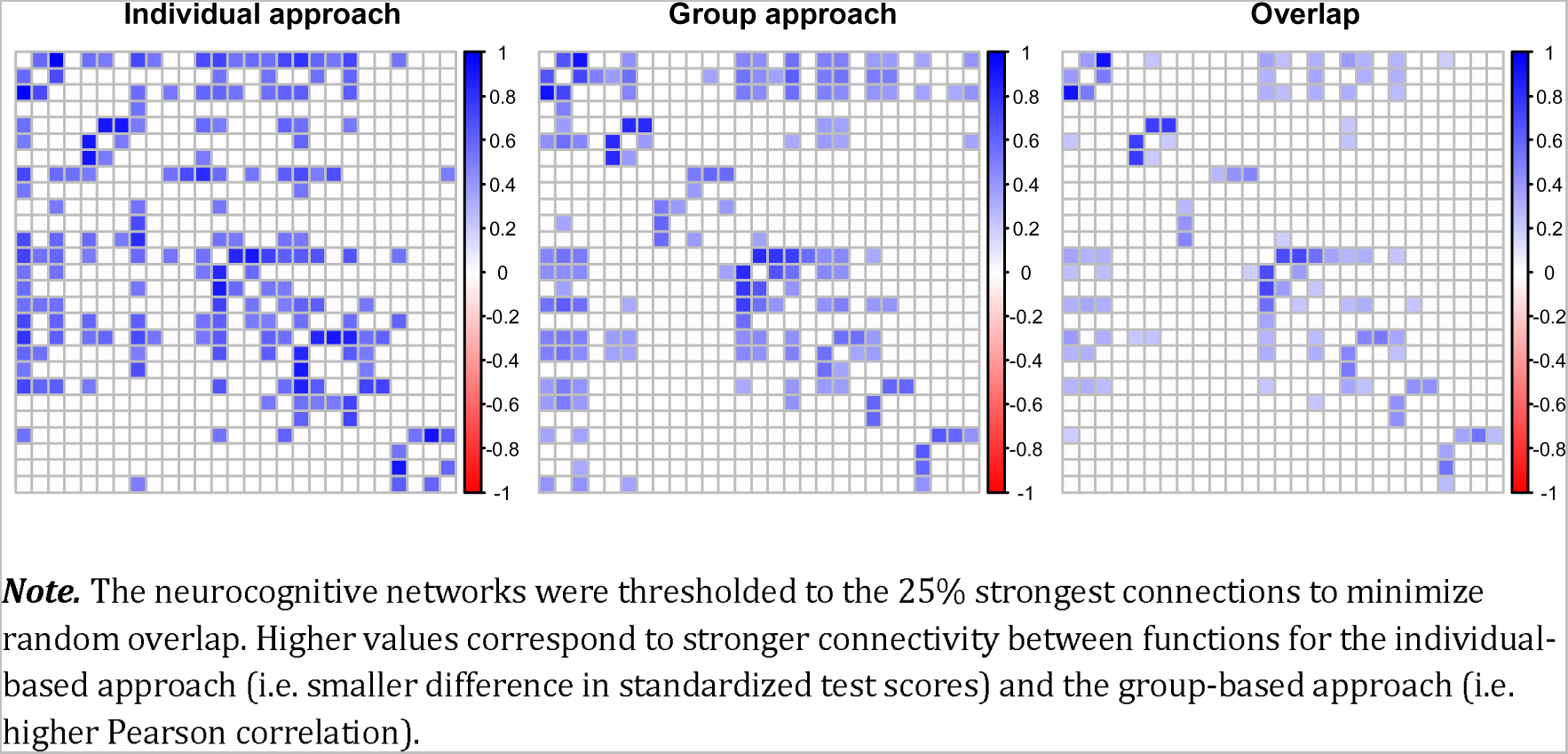
Correspondence between individual-based and group-based neurocognitive networks.

### 3.4 Stability of the individual network approach

The stability of the individual network approach was assessed by the consistency of the resulting neurocognitive network across two randomly selected subgroups of individuals (subgroup 1: n = 66 vs. subgroup 2: n = 66). Accordingly, we reconstructed the neurocognitive network in two randomly selected study subgroups and determined the overlap between the resulting neurocognitive networks (Figure S4). The results show that the networks from the two subgroups have considerable overlap, sharing 64.3% of their connections (bootstrap 95%-CI: 52.8% −75.8%). Again, the observed overlap was found to significantly exceed the overlap that would be expected on chance level (6.25%). This indicates that the individual network approach produces a robustly identifiable neurocognitive network across individuals. The results also reflect that there is considerable variability in the organization of the neurocognitive network, potentially reflecting relevant inter-individual variability in the configuration of neurocognitive functions.

**Figure S3.**
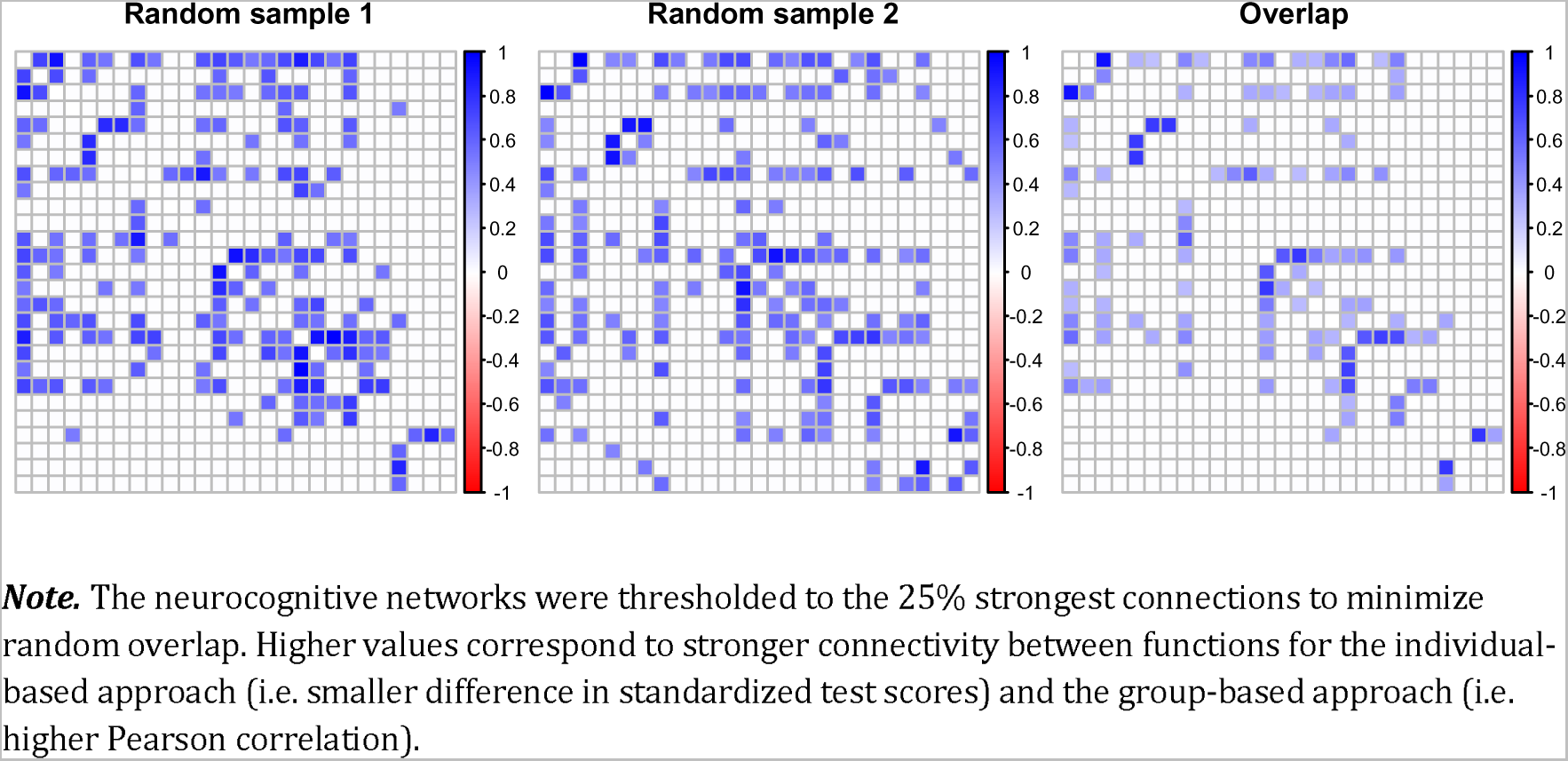
Correspondence between neurocognitive networks reconstructed in random samples.

### 3.5 Exploring global and local network organization

A graphical visualization of the neurocognitive network is provided in Figure S5. The organization of the neurocognitive network was explored at the level of the network as a whole (global network organization) and at the level of the process in the network (local network organization).

#### 3.5.1 Global network organization

Analyses aimed at exploring the influence of network threshold on global network organization (Figure 6) revealed effects for network threshold on all global network measures. As an expected consequence of increasing network threshold, average network strength decreased (F[1,10] = 27.5, p < .001), reflecting that sparser networks have a lower total connectivity value. Higher network threshold was also associated with higher assortativity (F[1,10] =658.4, p <.001). This finding reflects that sparser networks have more prominent hierarchy among neurocognitive functions in the network. Furthermore, we found that higher network threshold resulted in higher modularity (F[1,10] = 23.5, p < .001). This finding indicates that sparser networks have a higher degree of specialization, i.e. delineate into more subgroups of highly connected neurocognitive functions. Likewise, higher network threshold caused longer characteristic path length (F[1, 10] = 2660.0, p < .001), indicating that smaller networks have lower integration, i.e. a greater relative distance between neurocognitive functions. Higher network threshold had a negative impact on transitivity (F[1,10] = 362.9, p < .001), indicating that sparser networks have a lower level of clustering between neurocognitive functions. Smallworldness, representing the balance between clustering and integration in the network, was higher for higher network thresholds (F[1, 10] = 52.8, p < .001). This indicates that sparser networks tend to favor clustering over integration as compared to richer networks. Smallworldness was higher than one across the whole range of thresholds, indicating that the neurocognitive network typically has a small-world organization.

**Figure S4.**
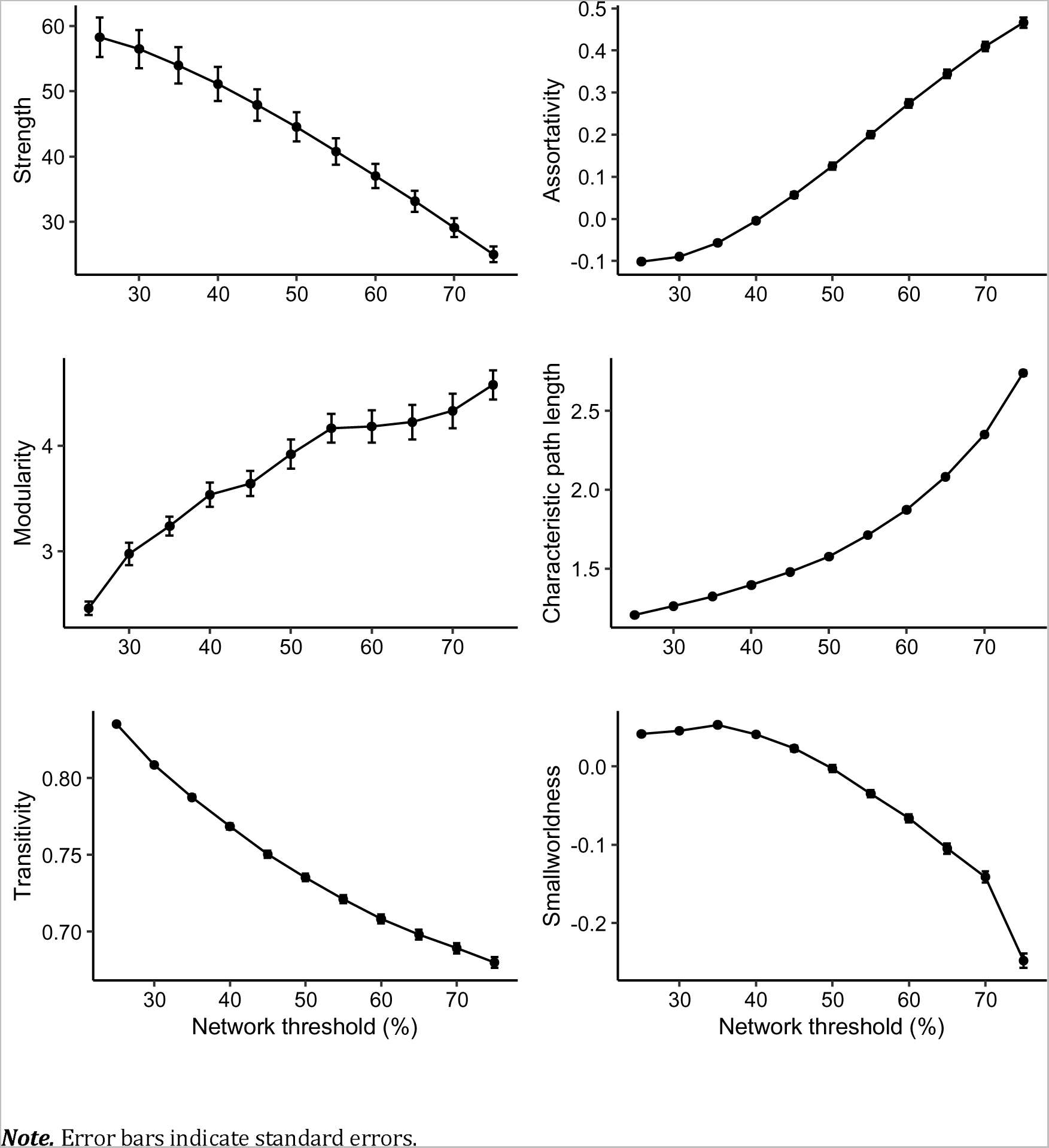
Global network organization across network thresholds.

**Figure S5.**
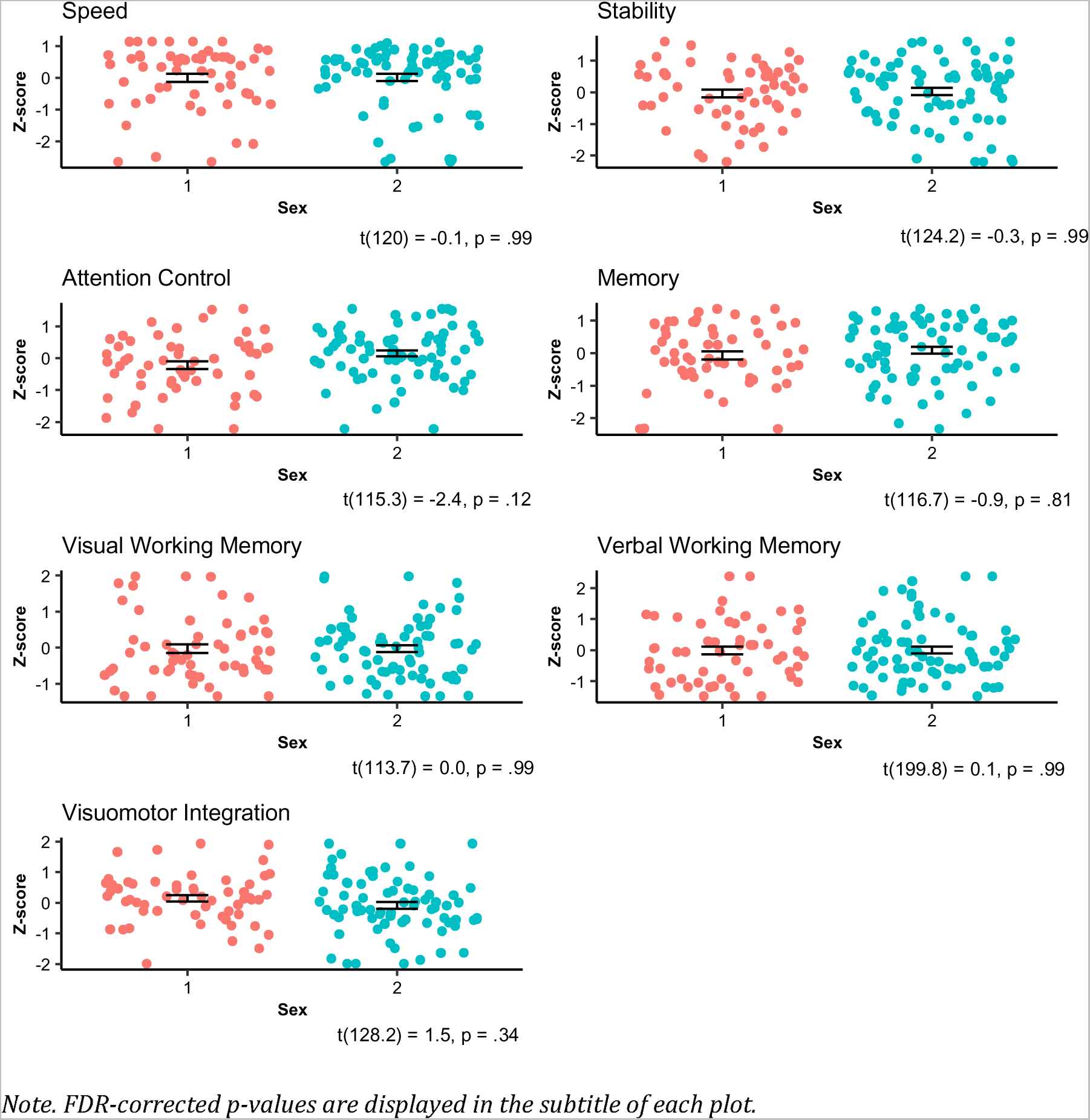
Sex and conventional neurocognitive domain measures.

**Figure S6.**
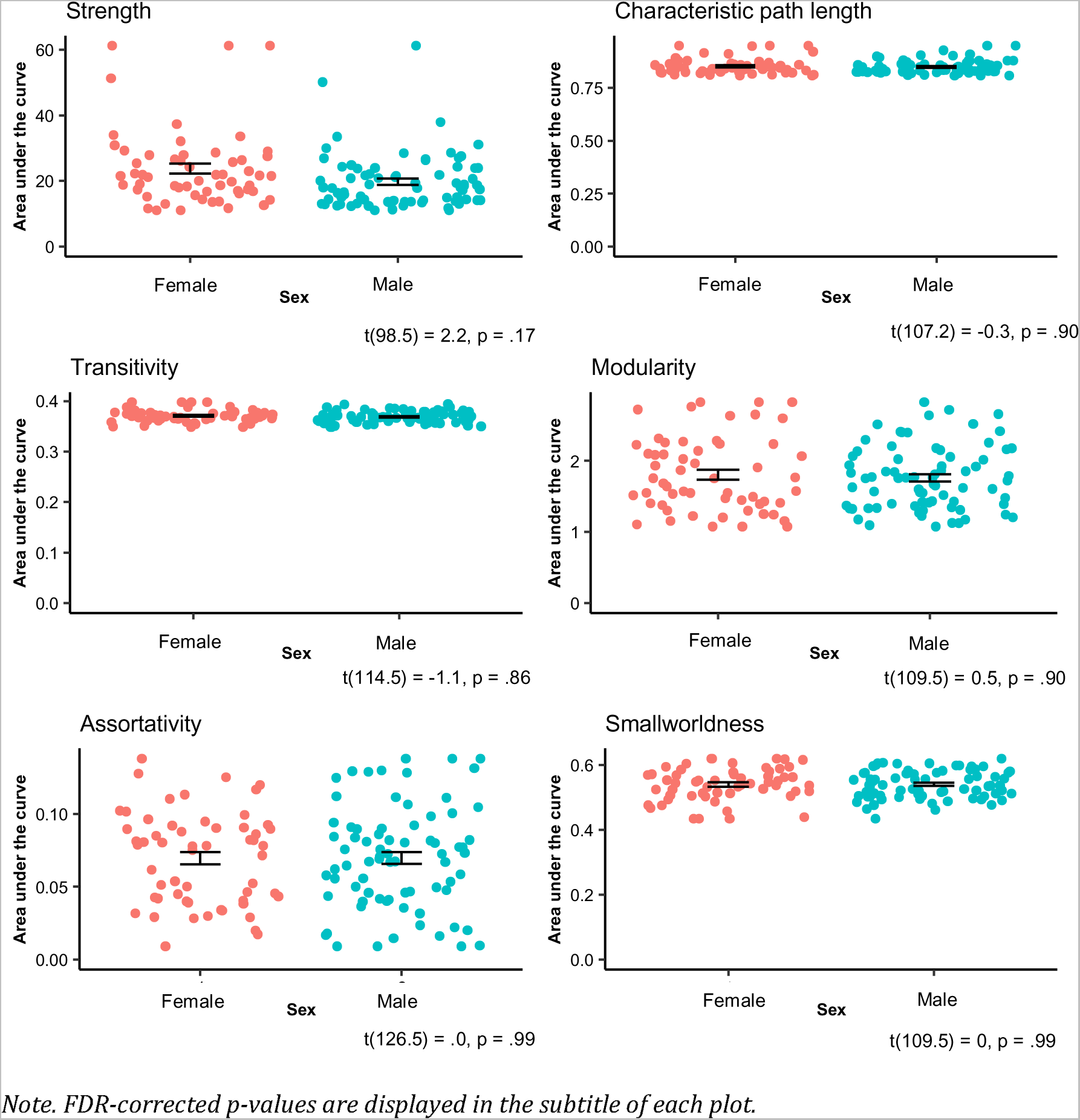
Sex and global network organization.

**Figure S7.**
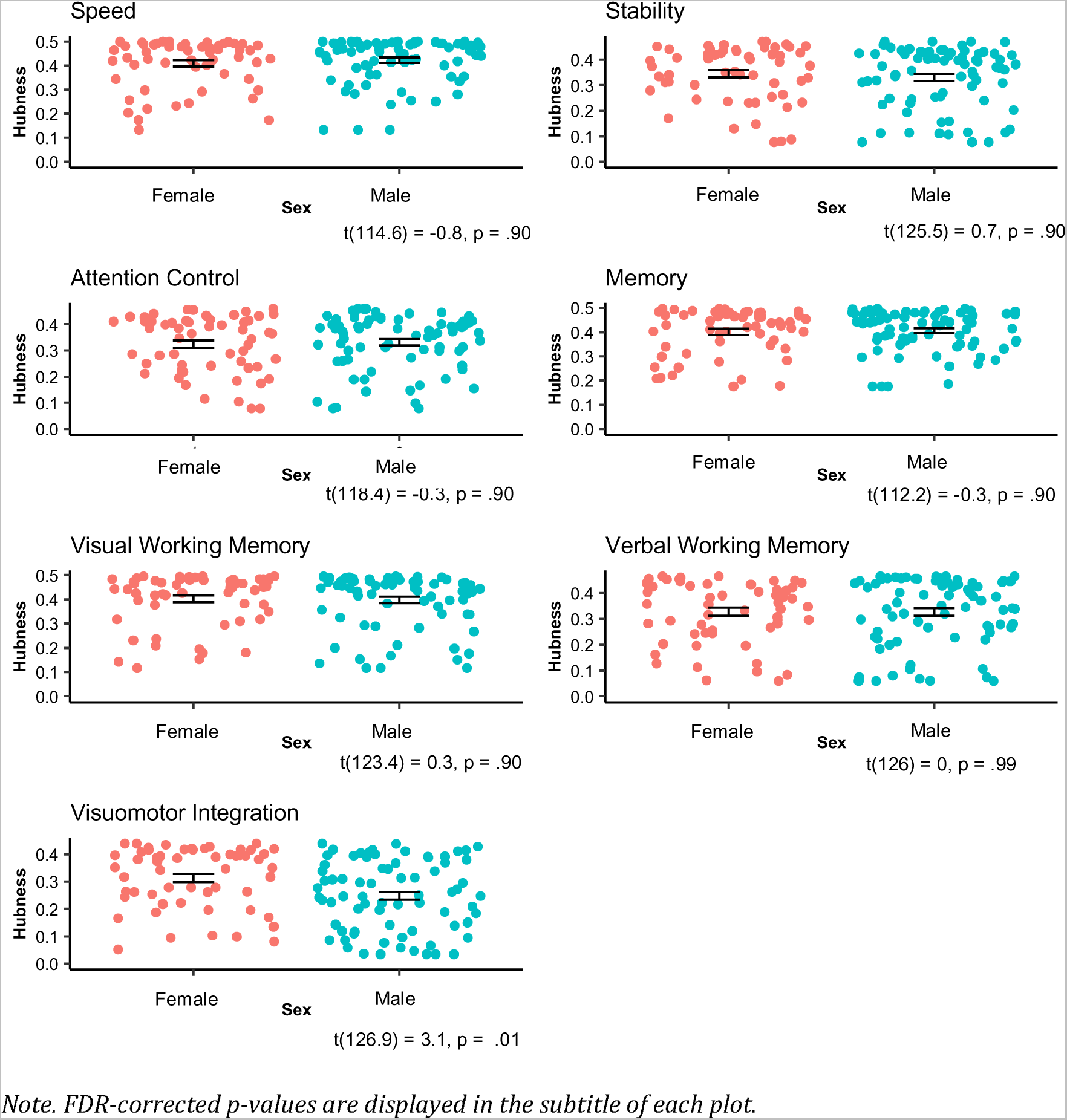
Sex and local network organization.

**Figure S8.**
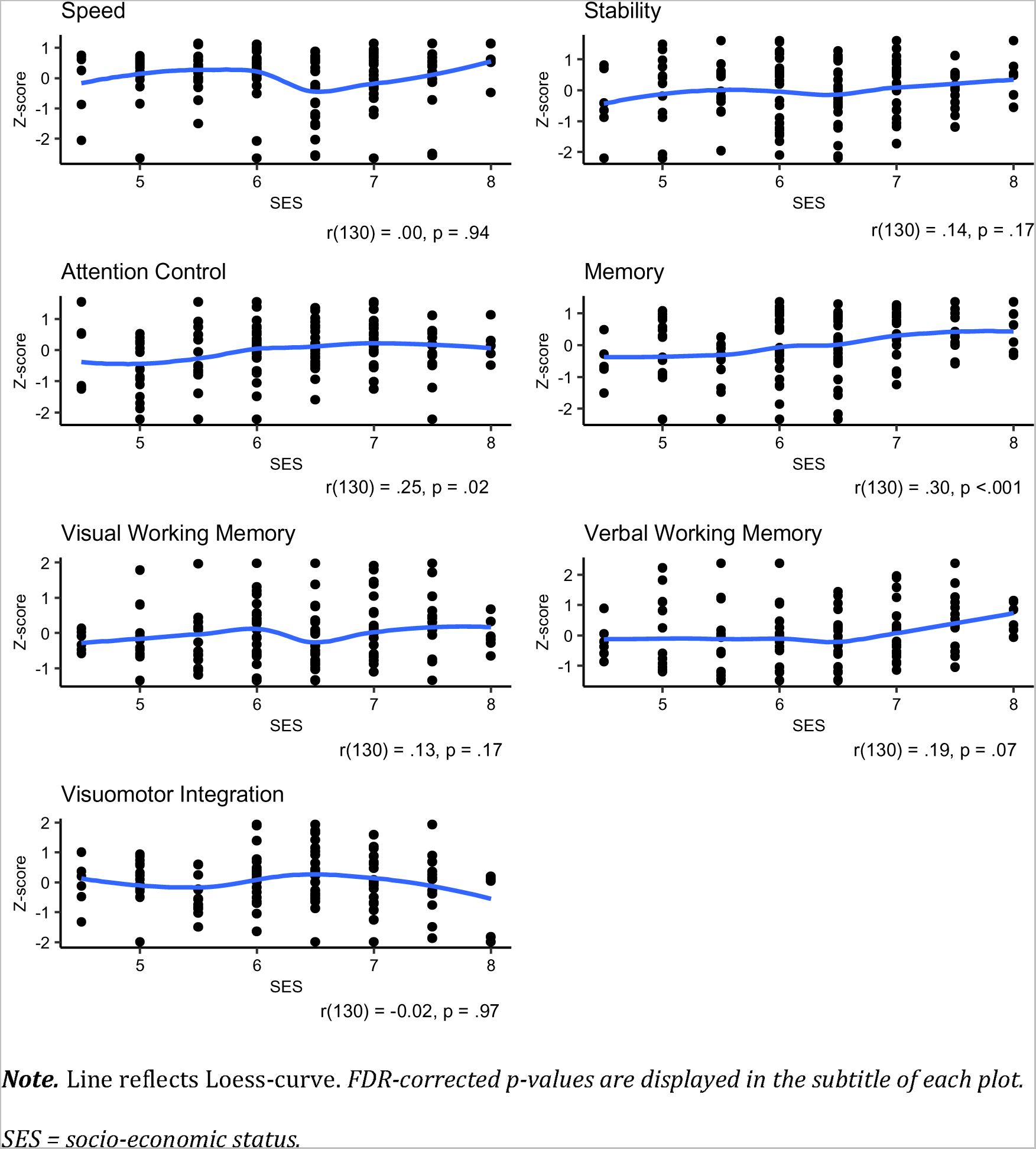
SES and conventional network organization.

**Figure S9.**
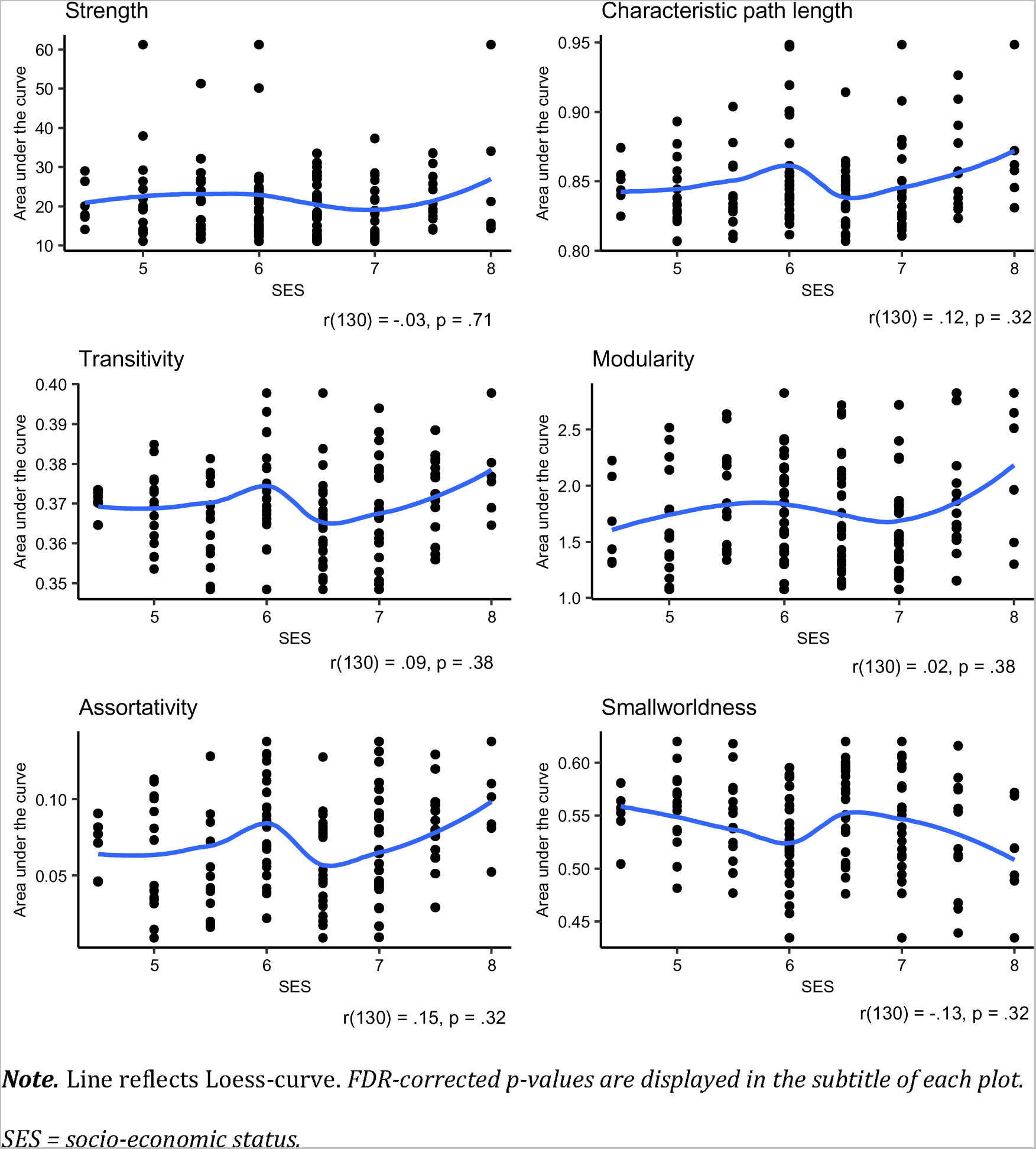
SES and global network organization.

**Figure S10.**
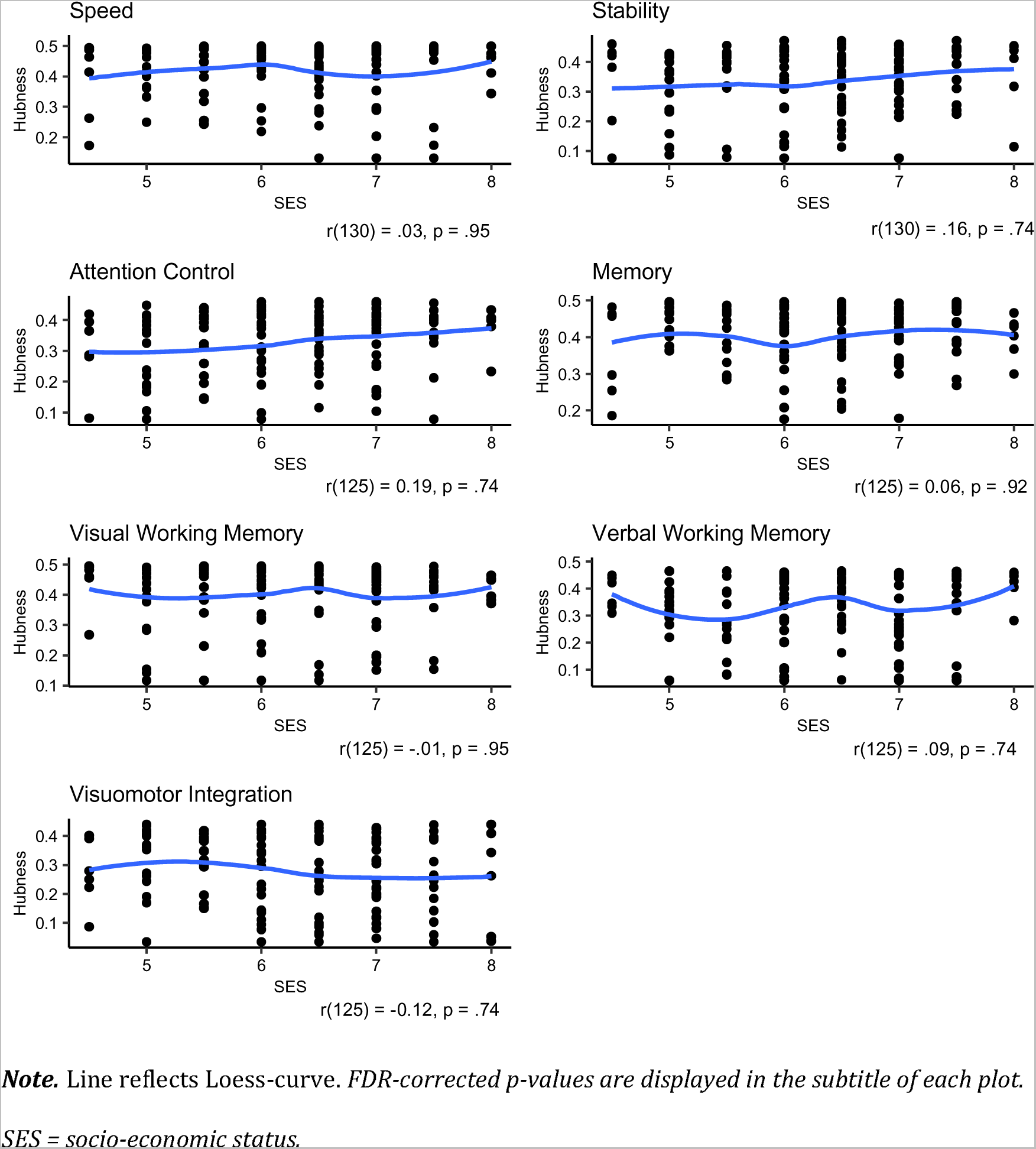
SES and local network organization.\

## Funding source

This work was supported by the ZonMw Offroad Program [grant number 04510011910052].

## Notes

### Competing Interest Statement

The authors have declared no competing interest.

### Author Declarations

Ethical approval was given by the medical ethical committee of the Amsterdam University Medical Centers (location AMC, NL76915.018.21).

